# On real-time calibrated prediction for complex model-based decision support in pandemics: Part 2

**DOI:** 10.1101/2025.05.16.25327744

**Authors:** Trevelyan J. McKinley, Daniel B. Williamson, Xiaoyu Xiong, James M. Salter, Robert Challen, Leon Danon, Benjamin D. Youngman, Doug McNeall

## Abstract

Calibration of complex stochastic infectious disease models is challenging. These often have high-dimensional input and output spaces, with the models exhibiting complex, non-linear dynamics. Coupled with a paucity of necessary data, this results in a large number of non-ignorable hidden states that must be handled by the inference routine. Likelihood-based approaches to this missing data problem are very flexible, but challenging to scale, due to having to monitor and update these hidden states. Methods based on simulating the hidden states directly from the model-of-interest have an advantage that they are often more straightforward to code, and thus are easier to implement and adapt in real-time. However, these often require evaluating very large numbers of simulations, rendering them infeasible for many large-scale problems. We present a framework for using emulation-based methods to calibrate a large-scale, stochastic, age-structured, spatial meta-population model of COVID-19 transmission in England and Wales. By embedding a model discrepancy process into the simulation model, and combining this with particle filtering, we show that it is possible to calibrate complex models to high-dimensional data by emulating the log-likelihood surface instead of individual data points. The use of embedded model discrepancy also helps to alleviate other key challenges, such as the introduction of infection across space and time. We conclude with a discussion of major challenges remaining and key areas for future work.

## 1 Introduction

In Part 1 [Williamson et al., 2025] we highlighted the utility of using Uncertainty Quantification (UQ) methods to calibrate complex, computationally-intensive infectious disease models. We also discussed key challenges in implementing these methods in real-time, and introduced four main ideas to help alleviate some of these, which are:

1. Set up a structured model discrepancy (MD) process and embed this within the simulation pipeline. In the models dicussed here we specify an idealised zero mean noise process, that explicitly operates to adjust the *hidden states* of the model over-and-above the *simulator* (i.e. it is not simply additional observation error). The MD is structured such that the adjusted hidden states are epidemiologically valid (that is, states are consistent with each other and the observed data).
2. Use a data assimilation mechanism as a structured way to build in information from available data to constrain the space of plausible trajectories. We propose using particle filtering to do this, which also generates unbiased estimates of the likelihood, which are typically more precise than naïve Monte Carlo estimators. This is fundamentally important for stochastic models, especially when simulating across complex populations such as those involving spatial, network or meta-population structures, since there are a huge number of potential trajectories that could be realised across the structured population. Without the data assimilation, the probability of enough simulations being close to the observed data with a finite number of model runs can become vanishingly small even in a perfect model/parameter scenario. As such the data assimilation facilitates *much* more efficient inference, as well as improving both prediction and forecasting. The combination of the embedded MD and filtering also tackles several other key challenges, most notably that of seeding (i.e. the introduction of infections in the early stages of an outbreak), since the MD provides a means to capture the effects of mechanistic processes that are not directly implemented in the simulation model. Due to the idealised zero mean structure, if excessive MD adjustments are required to track the observed data for a given set of parameters, then this will result in lower likelihood estimates than for parameter sets where the simulator can track the observations without requiring a large amount of adjustment.
3. Exploit the use of fast surrogate models (*emulators*) as an efficient means of exploring the parameter space given that the simulator and associated particle filter are computationally intensive to evaluate. Combined with the technique of *history matching*, which utilises a metric known as an *implausibility measure* to remove poor parts of the input space, this provides a powerful and robust way to calibrate models that explicitly accounts for major sources of uncertainty.
4. Emulate the log-likelihood estimates that we obtain from the particle filters. By emulating only a single output, this circumvents a key challenge with standard emulation techniques that require separate emulators for each output we wish to calibrate to, which for high-dimensional output spaces could undermine any potential efficiency gains of emulation. To do this we introduce a novel implausibility measure designed specifically for this output.

A generic framework for building models with embedded model discrepancy processes is laid out in Part 1 [Williamson et al., 2025], and in this paper we illustrate how these ideas can be extended to work with more complex, much higher-dimensional systems. Furthermore, we illustrate how this framework can be exploited to help efficiently tackle some further challenges, such as helping to alleviate particle impoverishment. The latter is a particular issue with particle filters where particle weights can concentrate on a small number of particles, which upon resampling results in a lower diversity of particle trajectories [see e.g. Gilks and Berzuini, 2001, Doucet and Johansen, 2011]. The motivating example for this work is a spatially-explicit transmission model of COVID-19 infection, defined across a meta-population of Lower Tier Local Authority (LTLA) districts in England and Wales. This model was developed during the 2020–2021 pandemic because it could have provided useful insights into localised spread, and the impact of movement restrictions. However, it was not possible to calibrate this model effectively in real-time, which hindered its practical use and motivated this series of papers. There are further additional complexities that must also be accounted for, since the observed data on hospitalisations and deaths—described in Section 2—are given at different spatial resolutions, requiring extensions to both the model structure and filtering algorithms. In Section 3 we describe the model structure, embedded model discrepancy, and observation processes. In Section 4 we describe the bootstrap particle filter, including the extensions to help alleviate particle impoverishment. In Section 5 we discuss the results of a simulation study, as well as models fitted to the real data.

A final key challenge with empirical filtering methods is that we can only evaluate a *finite* number of particles, and this affects both the estimates of the filtering distributions and the overall estimate of the log-likelihood (the unbiasedness property of the likelihood being an asymptotic result for infinite particles). It is not uncommon to see applied particle filtering papers use hundreds, or even thousands of particles. However, complex models can severely limit the numbers of particles that can be evaluated, due to both the length of time taken to evaluate the filters and also the memory requirements to store each particle. Only being able to run a small number of particles affects the efficiency of the particle filters, in terms of both their ability to track the observed trajectories, but also by decreasing the precision of the resulting log-likelihood estimates. We discuss the impacts of these and other challenges, as well as avenues for future work in Sections 5 and 6.

## 2 Data

All data processing and visualisation was done in the R statistical language R Core Team [2022] principally using the `tidyverse` [Wickham et al., 2019], `patchwork` [Pedersen, 2022], `sf` [Pebesma, 2018] and `areal` [Prener and Revord, 2019] packages. For a full list of packages used, please see the Supplementary Materials. We fitted the model to publicly available data on deaths and hospitalisations. The data used in this paper are available at https://github.com/tjmckinley/uq4covid.github.io/blob/forPaper2/. Unfortunately many of the original source links for the data are no longer active, however we provide documentation detailing where the data were downloaded from through the GitHub link above.

The *incidence* of deaths within 28 days of a positive COVID-19 test (i.e. the number of new deaths at each time point) are available either by LTLA (338 areas), or by age-class (*<*5, 5–17, 18–29, 30–39, 40–49, 50–59, 60–69, 70+ years) and region (‘East Midlands’, ‘East of England’, ‘London’, ‘North East’, ‘North West’, ‘South East’, ‘South West’, ‘West Midlands’ and ‘Yorkshire and the Humber’). Unfortunately, to our knowledge, there are no publicly available data stratified by age-class and LTLA. LTLAs are nested within regions, and lookup tables were found on https://geoportal.statistics.gov.uk and provided in the GitHub repository above. The data only provide total observed deaths and cannot delineate deaths in hospital from deaths in the community.

Hospitalisation data are provided at yet different aggregations. Hospital *incidence* data (i.e. the number of new patients with COVID-19 admitted to hospital) are available by age-class (0–5, 6–17, 18–64, 65+ years) and NHS-region (‘East of England’, ‘London’, ‘Midlands’, ‘North East and Yorkshire’, ‘North West’, ‘South East’ and ‘South West’). Hospital *case* data (i.e. the number of people in hospital with COVID-19 at a given time) are only available by NHS-region. For the purposes of calibration, counts at the LTLA level can be aggregated to region using the lookup tables described above, and can then be mapped to the corresponding NHS-region, assuming that both ‘East Midlands’ and ‘West Midlands’ map to the ‘Midlands’ NHS region, and that both ‘North East’ and ‘Yorkshire and the Humber’ map to ‘North East and Yorkshire’. Although not perfect, for the age to NHS-age categories we map: *<*5 → 0–5; 5–17 → 6–17; (18–29, 30–39, 40–49, 50–59) → 18–64; and (60–69, 70+) → 65+ years.

The movement data at the electoral ward level are provided in the MetaWards package [Woods et al., 2022], but were mapped from the original 2011 wards to 2019 wards using areal interpolation [Prener and Revord, 2019]. We then aggregate movements from the electoral ward level (*>* 8, 000 areas) to the LTLA level (338 areas), which is feasible since wards are geographically nested within LTLAs. Specific movements of groups of people between each pair of spatial locations is known as a *movement cohort*. Note that the movement (commuter) data includes wards in Wales, but the death and hospitalisation data only includes LTLAs and regions in England, and so although the underlying model simulates transmission within and between LTLAs in England and Wales, we only calibrate using simulated counts and data from England. Since the commuter data does not delineate by age, we split the population in each LTLA into age-categories according to the proportions of people in each age-class in the overall population, and then did the same for each movement cohort. These age-breakdowns were derived from publicly available records from the 2011 census licensed under the Open Government Licence v3.0 [ONS-NRS-NISRA, 2009].

## 3 Model structure

For ease-of-reference the specific model structure for COVID-19, introduced in Part 1 [Williamson et al., 2025], is reproduced in Figure 1. Briefly, all individuals begin in the *S* (susceptible) state. Transmission occurs through contact with individuals in any of the infectious states (*A, P, I*_1_ or *I*_2_), and on infection they enter the exposed (*E*) state, where they are infected but not yet infectious. Individuals will then progress through either the *asymptomatic* pathway, where they will eventually recover (*A* → *R*_*A*_), or they progress through the *symptomatic* pathway, passing first through a pre-symptomatic state *P*, and then onto a symptomatic state *I*_1_. Some symptomatic individuals will remain in the general population and eventually recover (*I*_2_ → *R*_*I*_), some will die without going to hospital (*D*_*I*_), and some will be hospitalised (*H*). Some hospitalised individuals will eventually die (*D*_*H*_), whilst some will eventually recover (*R*_*H*_). We assume that all recovered individuals have immunity to reinfection at the current time (since we are only modelling the short-term dynamics here). The model is specified in discrete-time over daily time intervals, and the probabilities of transitioning along different pathways are dependent on age, such that older individuals are more likely to transition down more severe pathways.

**Figure 1:**
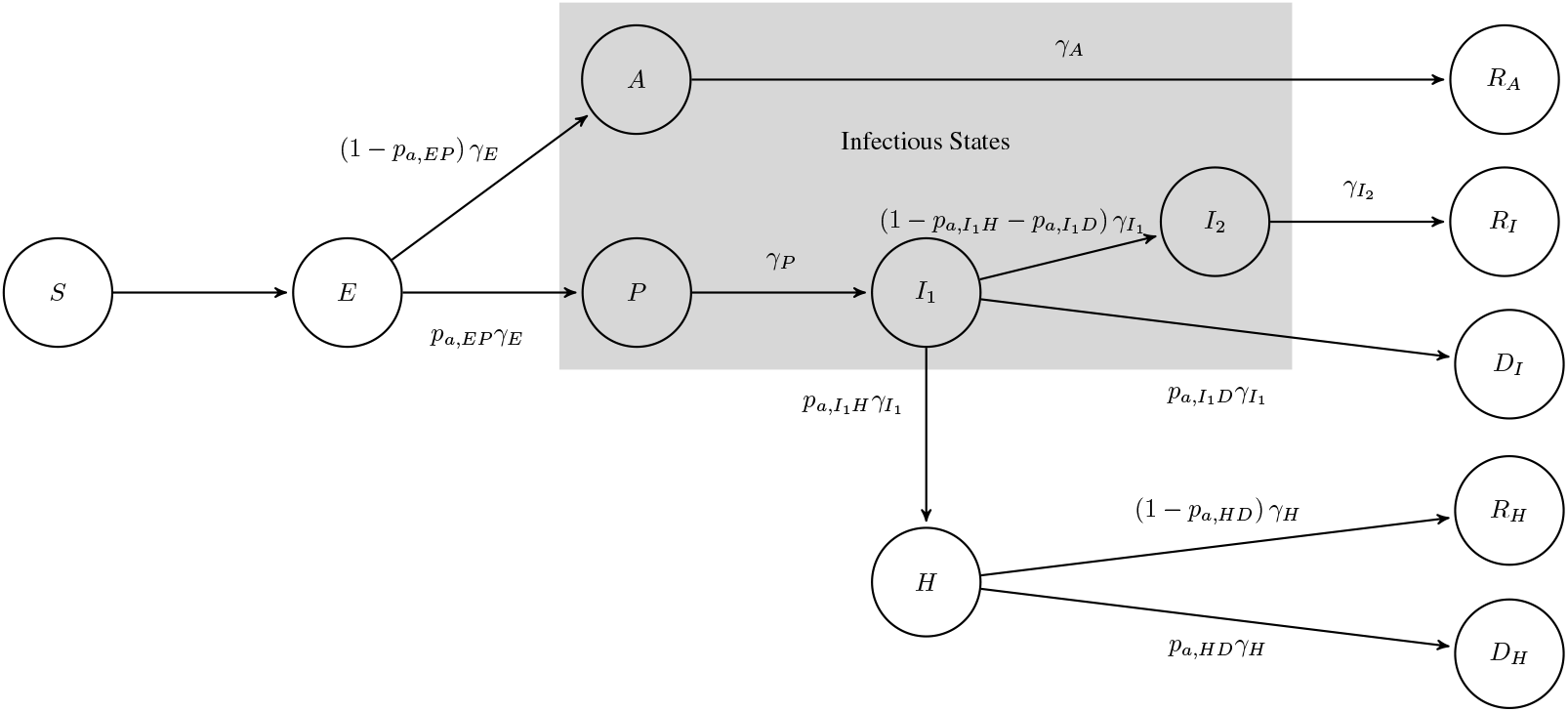
Schematic of the COVID-19 model considered in this paper and an example of the compartmental structure we consider throughout.

### 3.1 Notation

We use *X* to denote counts arising from the *transmission model, Y* to denote counts adjusted for the model discrepancy process, and *Z* to denote *observed* counts. We use primes to denote *incidence* counts (e.g. *X*^*′*^, *Y* ^*′*^, *Z*^*′*^), and subscripts to denote specific states, time points, age-classes and spatial areas. We assume that we have *N*_*a*_ age-classes, *N*_*s*_ spatial areas, *N*_*r*_ regions, *N*_*b*_ NHS age-classes, *N*_*n*_ NHS-regions and *N*_*t*_ time points.

At time *t*, we let ***X***_*t*_ = {***X***_*tas*_; *a* = 1, …, *N*_*a*_ and *s* = 1, …, *N*_*s*_}, where

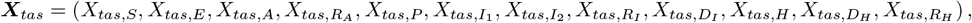

is a vector of counts from the *transmission model* at time *t* for age-class *a* in spatial area *s*. The transmission model structure is described in Section 3.2. Similarly, we let ***Y***_*t*_ = {***Y***_*tas*_; *a* = 1, …, *N*_*a*_ and *s* = 1, …, *N*_*s*_}, where

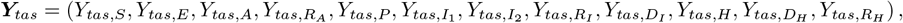

is a vector of counts *adjusted for model discrepancy* (MD) at time *t* for age-class *a* in spatial area *s*. The MD process is described in Section 3.3.

As described in Section 2, the observed data are not available at the same spatial resolution as the simulator and model discrepancy processes, and as such we let 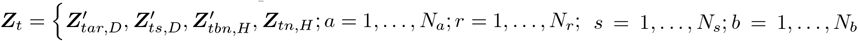 and *n* = 1, …, *N*_*n*_. Here the *D* state corresponds to the sum of *D*_*I*_ and *D*_*H*_. Section 3.4 describes how we map the MD-adjusted states ***Y***_*t*_ to the observed states ***Z***_*t*_.

In the discussion below we use *p* (·) to denote an arbitrary probability *mass* function. To be consistent with much of the particle filtering literature, we will use *g* (·) to denote a generic observation distribution across all observations, and *f* (·) to denote a joint state transition distribution.

### 3.2 Transmission model

Whenever we see forks in the pathways in Figure 1 we have multinomial transitions, otherwise we have binomial transitions. Hence the transitions between states in the period [*t* − 1, *t*) can be written (using · to denote the number of individuals who *do not transition* in the multinomial draws) as:

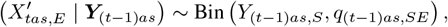

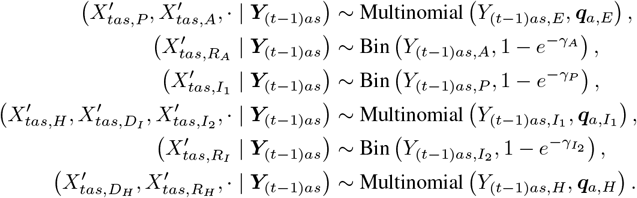

Here:

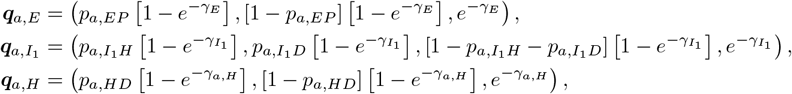

where

- *q*_(*t*−1)*as,SE*_ is driven by the number of infectious individuals in age-group *a*, area *s* at time *t* − 1, and a probability of transmission per infectious contact, *ν* (*R*_0_), which is defined as a function of the basic reproduction number, *R*_0_, and the next-generation matrix (NGM). These are discussed in more detail in Section 3.2.1.
- The transition rate 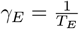 with *T*_*E*_ the mean latent period.
- The quantity *p*_*a,EP*_ is the probability that an individual in age-class *a* progresses through the *asymptomatic* pathway. We model log (*p*_*a,EP*_) = *α*_*EP*_ + *η* age_*a*_, where age_*a*_ is the mid-point of age-class *a*.
- The transition rate 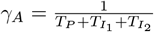 with 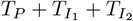 the mean infectious period.
- The transition rate 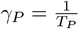 with *T*_*P*_ the mean pre-symptomatic infectious period.
- The transition rate 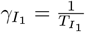 with 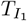 the mean pre-hospitalisation symptomatic infectious period.
- The probability that an individual in age-class *a* that is currently in the *I*_1_ state transitions down the hospitalisation pathway is given by 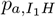, and the probability that they transition along the death pathway is given by 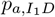. Similarly to before we model: 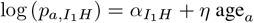 age_*a*_ and 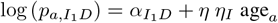, where *η*_*I*_ scales the age-relationship for *I*_1_ → *D*_*I*_ transitions.
- The transition rate 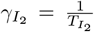 where 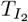 is the additional mean symptomatic infectious period for non-hospitalised individuals.
- The transition rate 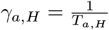 with *T*_*a,H*_ the mean length of hospital stay for an individual in age-class *a*. The length of hospital stays is age-specific, and is governed by the model log 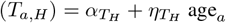.
- The probability that an individual in age-class *a* that is currently in the *H* state transitions along the death pathway is given by *p*_*a,HD*_. Similarly to before we model: log (*p*_*a,HD*_) = *α*_*HD*_ + *η η*_*H*_ age_*a*_, where *η*_*H*_ scales the age-relationship for *H* → *D*_*H*_ transitions.

#### 3.2.1 Transmission terms

Transmission is governed by the basic reproduction number, *R*_0_, which is defined as the expected number of secondary cases caused by a single infectious case introduced into a fully susceptible population. We also introduce a parameter, *ν*_*A*_, which allows asymptomatic individuals to have a different transmission potential to symptomatic individuals. The *force-of-infection* (FOI) acting on individuals in age-class *a* in area *s* at time *t, λ*_*tas*_, is therefore defined as:

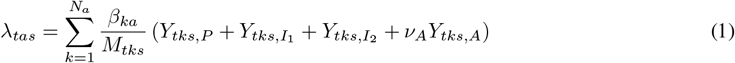

where *M*_*tks*_ is the population size of age-class *k* in area *s* at time *t*. Hence

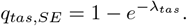

We can specify:

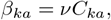

where 0 *< ν <* 1 is a probability of infection per contact, and *C*_*ka*_ is the *population contact rate* that an individual in state *a* has with individuals in state *k*. We use the population contact matrix *C* = {*C*_*ka*_; *k* = 1, …, *N*_*a*_; *a* = 1, …, *N*_*a*_} from the POLYMOD survey [Mossong et al., 2008] in the early stages of the outbreak, and then the CoMix survey after the first lockdown [Jarvis et al., 2020].

A key challenge is that since we parameterise our model in terms of *R*_0_, not *ν*, we thus need a way to derive *ν* for any given set of parameters. To this end, *R*_0_ can be derived as the *maximum eigenvalue* of the NGM [Diekmann et al., 1990, van den Driessche and Watmough, 2002, Diekmann et al., 2010, van den Driessche, 2017], and we can exploit this relationship here to derive *ν* as a function of *R*_0_, which we denote as *ν* (*R*_0_) here. For full details please see Supplementary Materials S2. We compute *ν* (*R*_0_) assuming the NGM defined in a single population of size 56,082,077 (matching the number of people in the commuter data) with 100 initial introductions at time 0. We note that the output is not sensitive to the exact number of susceptibles and infectives at time 0 as long as the number of susceptibles is large and the number of infectives is small. We also note that this mapping of *R*_0_ to *ν* is based on large population deterministic approximations that ignore the stochastic meta-population dynamics. As such the value of *R*_0_ that we calibrate to might not be a perfect representation of the *R*_0_ of our actual model, but it gives us a place to start (and it remains unclear how one would calculate *R*_0_ for a spatio-temporal model of this kind without making some of these simplifying assumptions).

#### 3.2.2 Movement cohorts

We use the framework set out in Danon et al. [2009] to model the impact of movements of individuals between spatial regions on the outbreak dynamics. The movement data has two components: *work* and *play* movements. Work movements consist of fixed moves of individuals between different LTLAs for the purposes of work. Conversely, play movements are governed by a stochastic process, where the numbers of individuals moving between specific LTLAs are governed by a series of multinomial distributions. Following the structure used in the MetaWards package [Danon et al., 2009, Woods et al., 2022], we assume that movements happen only during the day, and then all individuals return to their home LTLAs at night.

The transmission process is then applied twice at each time point, once during the day and once at night. As such the force-of-infection and the number of susceptibles in a given LTLA differs between the day and night due to movements. Furthermore we scale the FOI in each LTLA by 0.7 during the day, and 0.3 at night to reflect the relative lengths of the day and night. To ensure that individuals are moved correctly we keep track of the number of individuals in each state in each *movement cohort*, defined as a group of individuals who move between a specific pair of LTLAs. The number of movement cohorts is therefore much larger than the number of LTLAs, but from these we can reconstruct the numbers of susceptibles and infectives in each LTLA during both the day and night so that we apply the correct transmission process. Given a number of new infections in an LTLA, we can then redistribute these across the movement cohorts proportionately to the number of susceptibles in each cohort that maps to each given home LTLA (see Section 3.3.1).

All other transitions are assumed to happen at the end of the day, and we can simulate these within each movement cohort directly. This is because the probabilities of transition for individuals are fixed. As such the process of using binomial/multinomial sampling in each cohort and then summing these up within each LTLA is equivalent to sampling the transitions at the LTLA-level directly and then redistributing them proportionately to the relative counts.

### 3.3 Model discrepancy

For *absorbing states* (*D*_*H*_, *R*_*H*_, *D*_*I*_, *R*_*I*_ and *R*_*A*_) we place the model discrepancy (MD) on the *incidence* (i.e. new cases) rather than the state counts. For all other states, we place the MD on the numbers of individuals in each state directly. The MD terms are assumed conditionally independent within each age-class and area. In Part 1 [Williamson et al., 2025] we outlined an approach that ensures the MD process produces counts that are *epidemiologically valid*. For example, we can’t have more deaths than infections, or more hospitalisations than symptomatic cases and so on. This approach also ensures that the counts are truncated appropriately (e.g. we can’t have negative counts). As a result, although the MD is formulated as an idealised zero-mean noise process, in practice it is necessary to derive a series of ordered conditional discrete truncated distributions, where the discrepancy added to earlier epidemiological states is dependent on the discrepancy added to later epidemiological states. Note that we start by adding MD to the later states, and then progress back through to the earlier states to ensure validity.

In Part 1 [Williamson et al., 2025] we made a case that a natural choice for an MD distribution for the models/data under consideration here is the Skellam distribution, and we set up a framework for how to utilise and parameterise this distribution. The Skellam distribution can be approximated by a truncated discrete Gaussian distribution, which can be more computationally efficient. In this paper we decided to use truncated discrete Gaussian distribution throughout, and although we originally posited this as an approximation to the Skellam, it is perfectly reasonable simply as a direct alternative to the Skellam that has many of the same properties. Another motivation for this change was to explore ways in which the structure of the truncated discrete Gaussian might be leveraged to provide more efficient look-ahead particle filters, although we do not present any of that exploratory work here.

We denote a random variable *W* ∈ ℤ from a truncated discrete Gaussian distribution, with mean *µ*, variance *σ*^2^, lower bound *L* ∈ ℤ and upper bound *U* ∈ ℤ as:

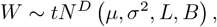

for −∞ *< µ <* ∞, *σ >* 0 and *L* ≤ *W* ≤ *U*. The probability mass function, and how to sample from this distribution efficiently, is described in the Supplementary Materials S1 for Part 1.

Therefore the MD for arbitrary state *c*, Δ_*tas,c*_, is specified as:

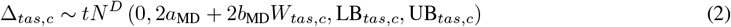

where *W*_*tas,c*_ corresponds to either the simulator incidence or count for the given state *c* (depending on which state the MD is being applied to) and parameters *a*_MD_ and *b*_MD_ allow the MD variance to increase as counts increase (see Part 1 Section 3.1). The full specification and derivation of the model discrepancy terms and bounds are given in Supplementary Materials Section S3.

#### 3.3.1 Redistribution across movement cohorts

Model discrepancy is applied at the LTLA-level, not the movement cohort level, and we apply the model discrepancy within each LTLA at the end of the day when all individuals are in their home LTLAs. However, since the transmission model is applied at the movement cohort level, we require a mechanism to redistribute the model discrepancy adjustments sampled at the LTLA level across the relevant movement cohorts. We do this proportionately to the numbers of affected transitions across all movement cohorts within each LTLA. In the following discussion, let ℳ_*tas*_ denote the set of movement cohorts at time *t* in age-class *a* that are nested in spatial area *s*. With a slight abuse of notation, we use the specific subscripts (*s* and *m*) to denote the aggregation level of the observation when referring to the spatial component. Since we simulate at the movement cohort level we also have access to ***X***_*tam*_ for all *m* ∈ ℳ_*tas*_ resulting from the simulator.

As an example of the redistribution process, consider that we wish to apply model discrepancy to hospital death incidence. Hence, once we have sampled 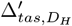 according to the process described in Section 3.3 and Supplementary Materials S3, then if 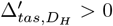 we must move additional individuals from the *H* state to the *D*_*H*_ state, whereas if 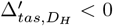 we will instead be reversing some of the *H* → *D*_*H*_ moves that have already occurred. An example algorithm for redistributing 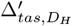 across movement cohorts *m* ∈ ℳ_*tas*_ is given in Algorithm 1. Similar algorithms can be derived for the other states in the model, but for brevity we have not shown these here.

#### 3.3.2 Spatially-varying model discrepancy initiation

We found it was necessary to add in some additional parameters that enabled the MD process to be phased in at different times in different spatial regions to reflect the fact that introductions of infections were not uniform across the country in the early stages of the outbreak. In each region *r* we allowed the MD parameters to vary over time, such that

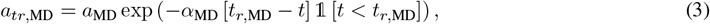

where 0 *< t*_*r*,MD_ *< t*^∗^, 0 *< α*_MD_ *<* 1 for *r* = 1, …, *N*_*r*_, and

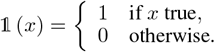

Hence, for *t > t*_*r*,MD_ then *a*_*tr*,MD_ = *a*_MD_, but for *t < t*_*r*,MD_ there is an exponential decay the further back in time you go from *t*_*r*,MD_, dependent on some rate parameter *α*_MD_. We also set

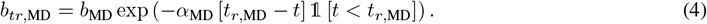

For all the examples in this paper, we chose *t*^∗^ to be equal to the time-of-first-lockdown, with *a*_MD_ = 0.05 and *b*_MD_ = 0.01. We decided to allow for greater model discrepancy variance within the oldest age-class, to represent additional key sources of uncertainty that were not included in the model, such as transmission in care homes. As such we set *b*_MD_ = 0.5 for the oldest age-class.

##### Algorithm 1 Redistribution of 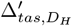 at time *t* for age-class *a* in spatial area *s* across movement cohorts *m* ∈ ℳ_*tas*_.

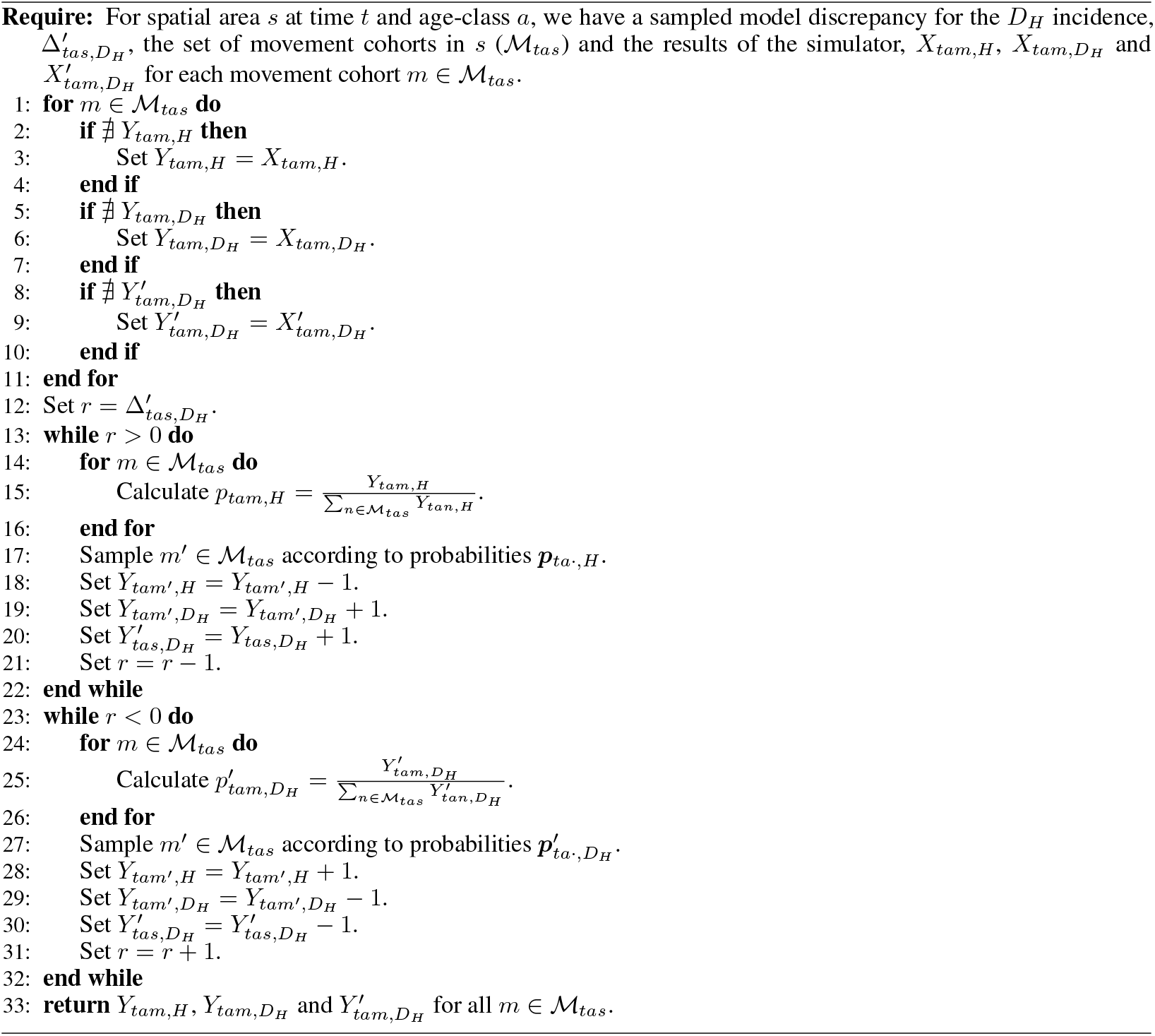

### 3.4 Observation distributions

A key challenge when using hospitalisation data is that the catchment areas of hospitals do not map cleanly to administrative boundaries, such as LTLAs. Various methods have been proposed to try to map NHS data to different spatial scales [e.g. Meakin et al., 2021, Challen et al., 2022], but this remains a considerable challenge. Given that we only have age-level data for deaths at the regional level, we decided to use a hierarchical observation process to map the LTLA/age-level information from the model to the higher-level aggregations in the data in a sensible way, allowing the observation process to reflect additional uncertainties in the mappings.

In the following discussion, let 𝒮 denote the set of LTLAs and 𝒜 be the set of age-classes used in the model. Also let ℛ_*r*_ ⊂ 𝒮 be the set of LTLAs that make up region *r* (*r* = 1, …, *N*_*r*_); 𝒩_*n*_ ⊂ 𝒮 be the set of LTLAs that make up NHS region *n* (*n* = 1, …, *N*_*n*_); and ℬ_*b*_ ⊂ 𝒜 be the set of age-classes that make up NHS age-class *b* (*b* = 1, …, *N*_*b*_). With a slight abuse of notation, we use the specific subscripts (*a, b, s* and *r*) to denote the aggregation level of the observation. Hence at each time *t* we observe the death incidence by age-class and region, 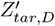 (*a* = 1, …, *N*_*a*_ and *r* = 1, …, *N*_*r*_), and the death incidence in each LTLA, 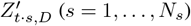. We also observe the hospital incidence by NHS age-class and NHS region, 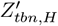(*b* = 1, …, *N*_*b*_ and *n* = 1, …, *N*_*n*_), and the number of people in hospital in each NHS region, *Z*_*t*·*n,H*_ (see Section 2 for more discussion). We know that the data at different aggregations are not independent; for example, 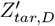 and 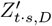 are correlated for *s* ∈ ℛ_*r*_. Furthermore, there are additional errors in the data sets which means they do not exactly match when aggregated up to a common level (for example, 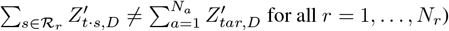.

One way to deal with these issues is to introduce a set of latent variables, denoting the hypothetical observation error (OE) at the LTLA/age-class level of the model. For example, for deaths we could introduce 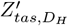 and 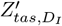, from which:

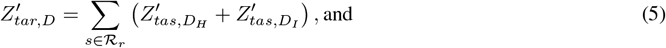

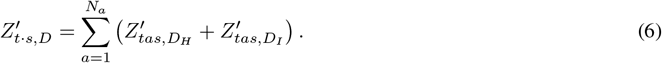

Given functional forms for the p.m.f.s of 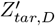 and 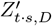, one may be able to derive the relevant joint distribution 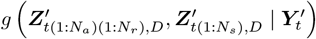. In Part 1 we discussed the use of Skellam distributions for the observation error, but as for the MD terms, here we chose truncated discrete Gaussian distributions instead. As such, we could use

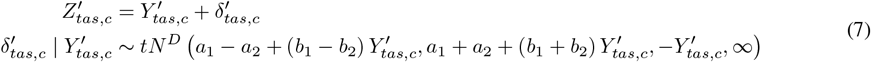

for the OE around the *incidence* for some state *c* (with analogous forms if the OE is placed around a *count* instead of the incidence). Here *a*_1_, *a*_2_, *b*_1_, *b*_2_ *>* 0 are parameters that enable us to control the mean and variance of the OE process and their dependence on the hidden states (see Part 1 Section 3.1.3).

We note that the upper bound of the OE should be finite (bounded by the data), but in practice the hidden states represent a small proportion of the overall population and unless the OE variance is very large, (7) is a suitable approximation. We set *a*_1_ = *a*_2_ = *a*_OE_ and *b*_1_ = *b*_2_ = *b*_OE_ here, which simplifies to

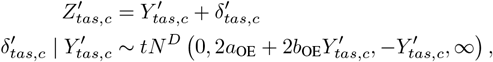

or equivalently

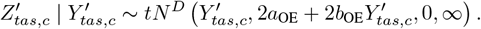

We note that setting *a*_1_ *< a*_2_ allows for average under-reporting to be introduced, but with the benefit that the OE will always have a non-zero probability mass function (in contrast to a stricter binomial-type OE model, which will have a zero likelihood if the hidden states are simulated to be larger than the observed states. This helps to alleviate some challenges with particle depletion in particle filtering methods). We will explore these variations in future work.

However, another challenge is that the sums of truncated discrete Gaussians in equations (5)–(6) do not have convenient mathematical forms, so instead we work with a set of *continuous, unbounded* latent variables,

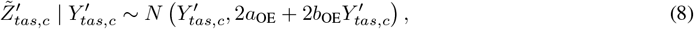

where 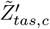 now corresponds to the *expected* observation given the hidden states. Since these are Gaussian, then any sum of these is also Gaussian. For deaths say, this allows us to specify:

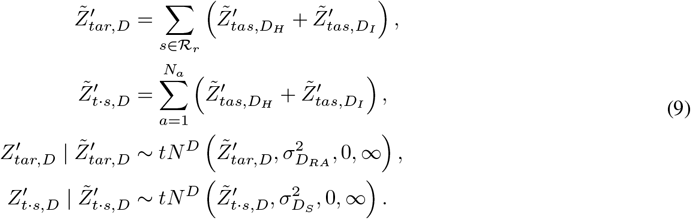

Here 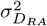 and 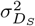 are additional variance terms chosen to represent additional aggregation errors at the regional/age-class and LTLA levels respectively (described in Section 2).

Since the OE for hospitalisations occurs on both the *incidence* and *counts*, they are not independent of the OE for deaths, since hospital case numbers depend on death and removal incidence as well as hospital incidence. As such, we also introduce latent variables for the expected observed hospital removals for each age/LTLA. Hence we can specify:

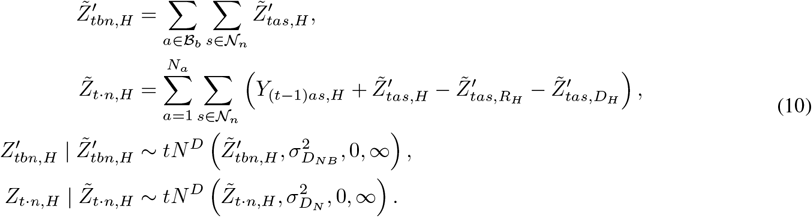

Here 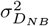 and 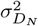 are additional variance terms chosen to represent the additional aggregation errors at the NHS region/NHS age-class and NHS region levels described above. The correlation structure between the multiple aggregations of death and hospitalisation data is thus captured through this hierarchical latent process. The aggregation error process captures the additional recording errors as described above. In the examples in this paper we chose *a*_OE_ = 0.001 and *b*_OE_ = 0.025. We then chose 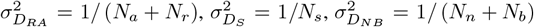 and 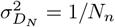.

### 3.5 Likelihood

To facilitate some of the later discussion regarding the particle filters, let’s consider the form of the likelihood functions for the simulator, model discrepancy and observation terms as described above. In all discussions below we drop dependence on the parameters, ***θ***, for brevity. Since we have fixed initial conditions, the target likelihood can be written as:

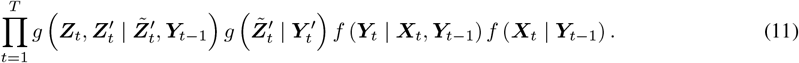

(The dependence on ***Y***_*t*−1_ in 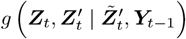 is required for calculating 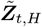 in (10).) The component relating to the transmission model can be written as:

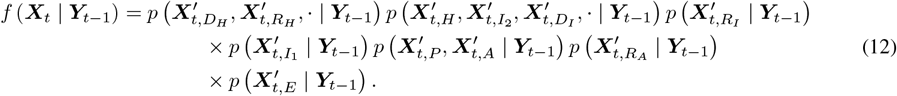

The infection distribution, 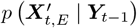 is a joint distribution over all spatial areas, age-classes and movement cohorts, whereas all other transitions are independent over area and age-class. For example:

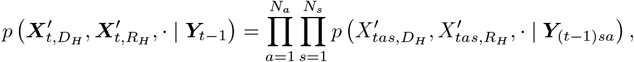

and similarly for the other transitions. See Section 3.2 for details of each transition distribution.

The conditional model discrepancy distribution can be written as:

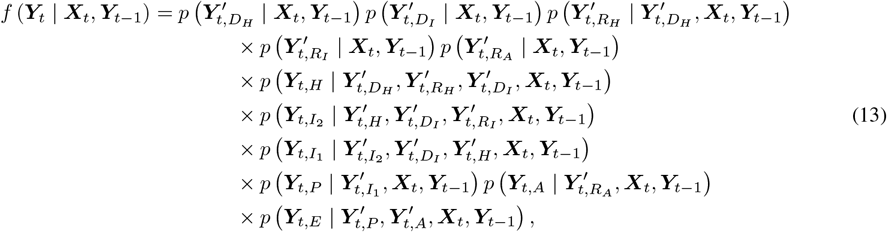

and here all the component distributions in (13) are independent over spatial area and age given ***X***_*t*_ and ***Y***_*t*−1_. We note that some of the terms depend on **incidence** counts from the transmission model 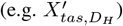, but these can be directly derived from ***X***_*t*_ and ***Y***_*t*−1_ as required. To keep notation as simple as possible, we have only made explicit the conditional dependence of the various MD states on other MD states in (13).

The first component of the conditional observation density can be written as:

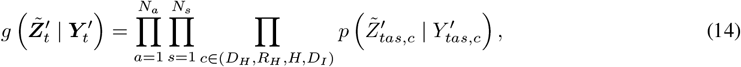

where the 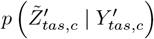 are given by (8). Then we have

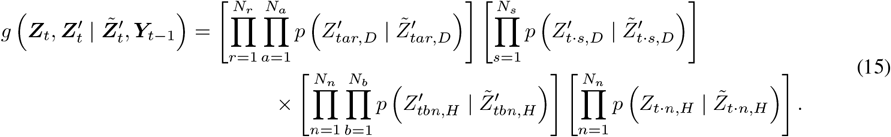

As an aside, we note that for other data structures it may be possible to analytically integrate across the continuous latent states, in which case the observation density could be written as:

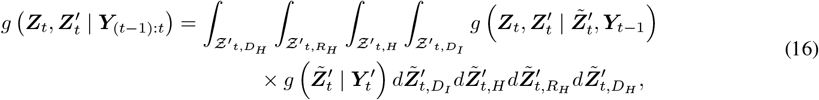

where the 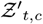 represent the multi-dimensional latent spaces for the 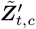 terms for state *c* ∈ (*D*_*H*_, *R*_*H*_, *H, D*_*I*_). If possible, this would reduce the uncertainty in the likelihood estimates from the particle filter caused by having to simulate the additional latent states.

## 4 Bootstrap particle filter

A standard bootstrap particle filter (BPF) [Gordon et al., 1993] for this model is shown in Algorithm 2, along with some amendments to help alleviate particle impoverishment using Markov chain Monte Carlo (MCMC, Gilks et al., 1996) steps [Gilks and Berzuini, 2001, Doucet and Johansen, 2011].

### 4.1 Tackling particle impoverishment

In a complex spatial meta-population model such as that described here, it is common (with a small, finite number of particles), to experience partial, or complete, particle degeneracy and impoverishment at any given time *t*, thus resulting in the particles having low diversity and in extreme cases all particle trajectories can collapse onto a single trajectory. Following e.g. Gilks and Berzuini [2001] and Doucet and Johansen [2011] we can introduce variability by taking each particle in turn and performing a series of *N*_iter_ MCMC steps, using a Markov kernel, *K*_*t*_(·), of invariant distribution *p* (***X***_1:*t*_, ***Y***_1:*t*_ | ***Z***_1:*t*_). Such a kernel is only ergodic if all ***X***_1:*t*_ and ***Y***_1:*t*_ are updated, but this would be too computationally intensive, and so following Doucet and Johansen [2011] we will update only **(*X***_*t*_, ***Y***_*t*_ | ***X***_1:(*t*−1)_, ***Y***_1:(*t*−1)_, ***Z***_1:*t*_)at each iteration *t*. We exploit the structure of the model and MD terms to produce efficient updates using *independence sampler* Metropolis-Hastings steps. The reader is referred to e.g. Gilks et al. [1996] for a more detailed introduction to MCMC.

To simplify future discussions, note that (from Section 3.2) we can decompose

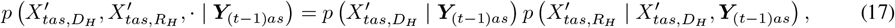

and

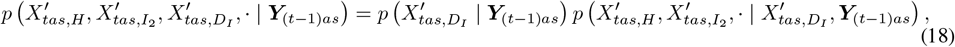

and since the distributions on the left-hand sides of (17) and (18) are multinomial, the marginal distributions on the right-hand sides are binomial and the conditional distributions are either binomial or multinomial.

Letting 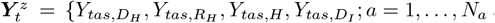 and *s* = 1, …, *N*_*s*_} be the adjusted states at the age-class/LTLA level that are necessary for mapping to the observed states ***Z***_*t*_, with 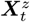 denoting the corresponding states generated from the simulator, then based on (11)–(18) we can write the joint likelihood for the hidden states and the data at time *t* as:

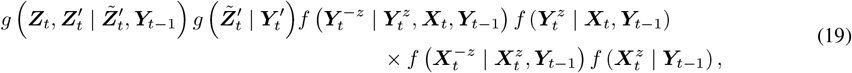

where we denote 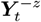 as those states in ***Y***_*t*_ that *do not* include those in 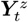, and similarly for 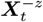.

We can further simplify the dependencies, and then exploit these to produce efficient update schemes. Firstly, from Section 3.3 we can see that

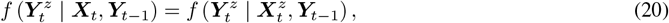

and from (14)–(15) we can also see that

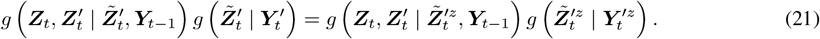

#### Algorithm 2 Bootstrap Particle Filter with MCMC steps

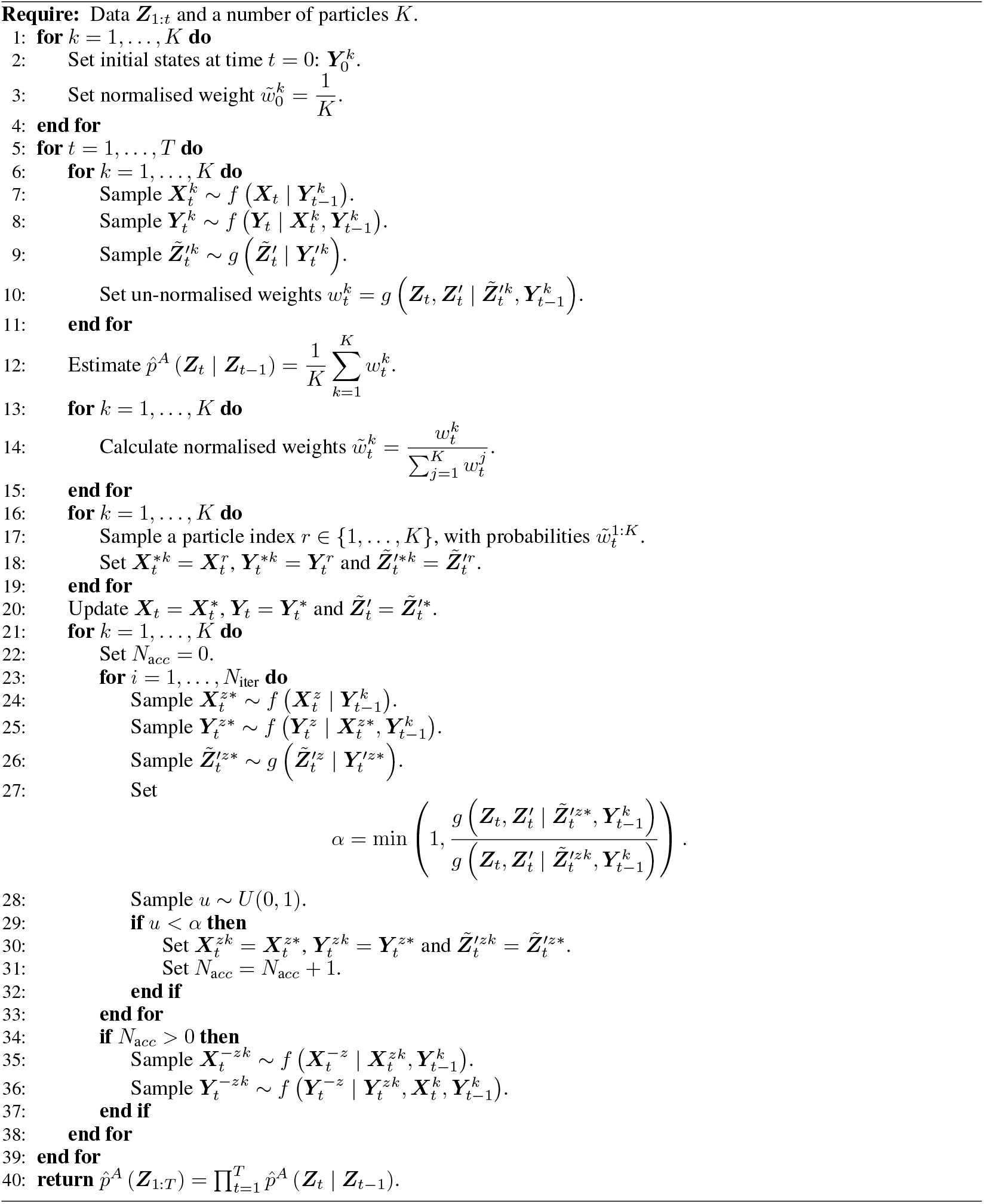

From the decomposition in (19) we can then write the likelihood contribution at time *t* as:

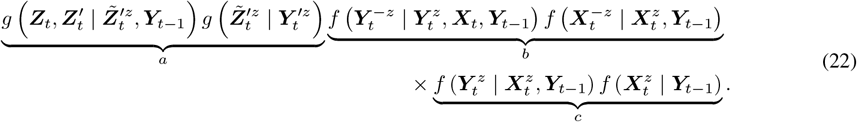

From (12) we have that:

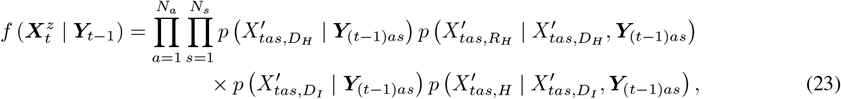

and from (13) and Section 3.3 we have that:

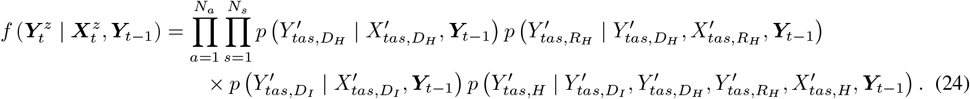

For a given particle *k*, we can simulate proposals 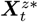 and 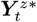 from (22c) easily by sampling directly from (23) and then (24). We can then simulate proposals 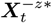 and 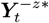 given 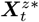 and 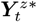 from (22b), by sampling directly from the conditional distributions 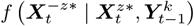 and 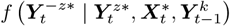 respectively. Details for how to sample from these conditional distributions are given in Supplementary Materials Section S4, and one particularly desirable feature is that 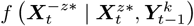 can be sampled from by simply changing some of the inputs in the simulator model, meaning that it can be evaluated with minimal changes to the code-base. Finally, we can sample proposals 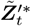 directly from 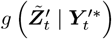 defined in (14).

With this proposal mechanism and the likelihood structure in (22), we can see that these proposals are accepted with probability:

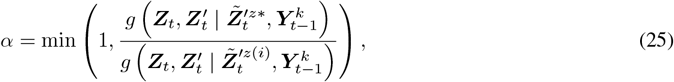

where 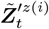 is the value of 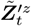 at iteration *i* (*i* = 1, …, *N*_iter_).

The computational burden of this proposed update scheme is sampling from (22b). However, it is worth noting that we do not actually have to sample from (22b) at all in order to evaluate the accept-reject ratio (25). Furthermore, using the proposal mechansim above means that 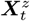 and 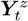 can be sampled *independently* of 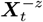 and 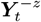 given 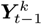 meaning that multiple iterations of these update steps can be done quickly and efficiently without having to sample from (22b). If, at the end of these iterations at least one proposal has been accepted, then we can do a single draw of 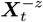 and 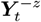 respectively, given the current values of 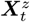 and 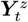. This is much more efficient than sampling from (22b) multiple times. Since proposals can be rejected, running multiple update steps increases the probability of accepting a proposal, and thus increases the likelihood of reducing particle impoverishment.

### 4.2 Initial plausible regions

Details of the derivations of the initial plausible regions for most parameters are given in Part 1 [Williamson et al., 2025]. In addition, for this spatial model we set prior ranges for the introduction of MD in each spatial region between the start of the study and the time of first lockdown (*t*_lockdown_). The parameter *p*_move_ (see Section 5.2.2) is bounded in (0, 1), and *α*_MD_ is bounded in (0.1, 1). The parameters *η*_*I*_ and *η*_*H*_ are allowed to vary between (0.5, 2).

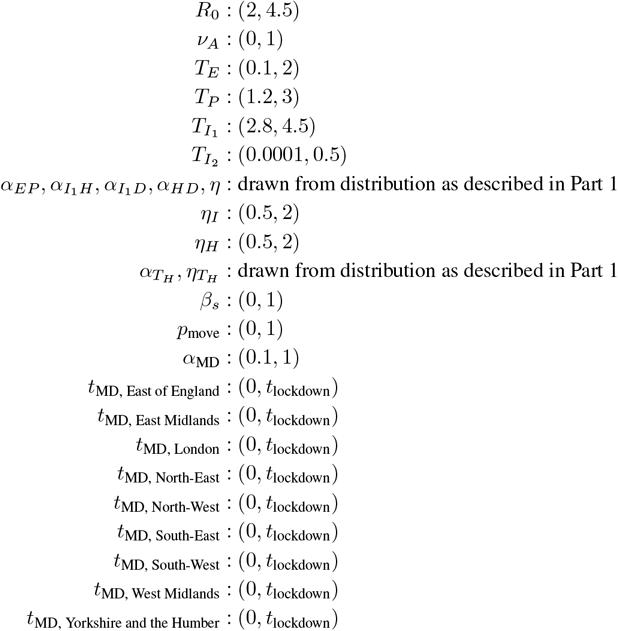

### 4.3 Model implementation

The meta-population framework described above and developed originally in Danon et al. [2009] is implemented in the open-source MetaWards software [Woods et al., 2022, available at https://metawards.org/]. Indeed originally we coded the simulator up in MetaWards, although currently this software does not natively support particle filtering, and as such we rewrote our specific model in R using the `Rcpp` [Eddelbuettel and François, 2011] and `RcppArmadillo` [Eddelbuettel and Sanderson, 2014] packages, which allowed us to implement the MD process and bootstrap particle filter developed here. For the emulators we used deep Gaussian Processes (DGPs), using the approach of Ming et al. [2023], implemented in the `dgpsi` package in R [Ming and Williamson, 2024]. History matching was performed using the hmer package in R [Iskauskas et al., 2024]. For a full list of packages used, please see the Supplementary Materials. All code used to run the models in this paper can be found at https://github.com/tjmckinley/uq4covid.github.io/blob/forPaper2/.

### 4.4 Emulation and history matching

For Wave 1 we generated an initial input design using Latin Hypercube Sampling for the uniform ranges, and then augmented this with a space-filling design over the non-uniform ranges as described in Part 1. For each wave we generated a set of 200 design points, and a further set of 50 validation points.

For each design point ***θ***_*i*_ we ran the BPF in Algorithm 2 and generated an estimate of the log-likelihood, *l* (***θ***_*i*_). Using dgpsi, we then trained a two-layer DGP emulator with a heteroscedastic Gaussian likelihood function on these runs, and validated the emulator using the hold-out validation runs. To reduce the complexity of the emulator, we selected a set of active variables using automatic relevance determination (ARD) where we fitted an initial standard GP, and then removed any input parameters with large length-scales. We chose length-scale thresholds for each wave by comparing the estimated length-scales with each other, and then checking that the validation plots were not significantly affected by the removal of the inactive variables. Then the DGP was fitted to just the active variables identified by the ARD process. If the fitted DGP passed the out-of-sample validation checks, then we augmented the training data with the validation data to produce a final updated emulator. We got much improved validation by emulating *h* (***θ***) = log [−*l* (***θ***)] instead of *l* (***θ***) directly.

As described in Part 1, we developed a novel implausibility measure to define the Not Ruled Out Yet (NROY) space, such that:

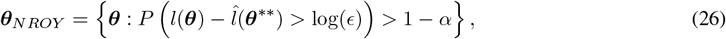

where 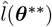 the current *maximum* log-likelihood estimate from the simulator runs. Since here we emulate *h* (***θ***), we note that

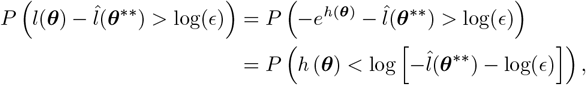

and thus we can amend (26) accordingly, such that

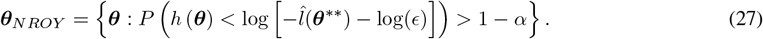

We set *α* = 0.95 with *ϵ* = 0.00001 throughout.

By definition, ***θ***^∗∗^ is in the tails of the emulated log-likelihood surface, and so on rare occasions the emulator will pass all usual diagnostics but rule-out ***θ***^∗∗^ if the emulator does not capture the tails of the log-likelihood surface well enough. To ensure that ***θ***^∗∗^ is always retained in the NROY space, we adapted the local Voronoi tessellation approach of Xu et al. [2021], where we retain regions around a set of ‘doubt points’, which are points that the emulator wants to rule out, but for which there is some evidence should be retained. To generate doubt points we first select all the training and validation points that the emulator wishes to rule out (using the implausibility measure based on the emulator in (27)). Then, for each *ruled-out* point we calculate a version of the implausibility based on the log-likelihood estimates from the particle filters directly, rather than through the emulator approximations (in some papers this would be known as the ‘simulator implausibility’—see e.g. Iskauskas et al., 2024). Doubt points (***θ***^*D*^) are then the subset of ruled-out points that would *not* be ruled out by the simulator implausibility, here defined as:

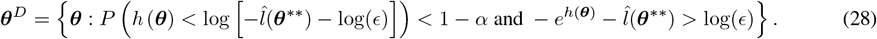

If ***θ***^*D*^ is empty, then we can proceed with our history matching procedure as normal. If ***θ***^*D*^ is not empty, then we further augment ***θ***^*D*^ by including any design point which is closer to a doubt point than any other design point, where distance is measured using the emulator posterior covariance function. This final set of augmented doubt points are automatically retained in the NROY space, but furthermore any new point that lies closer to a doubt point than any other design point, is automatically retained in the NROY space, where distance is once again defined using the emulator posterior covariance function.

Once we have defined the remaining NROY space in each wave, we used the slice-sampling approach of Andrianakis et al. [2017]—implemented in hmer—to sample from it. To generate a space-filling design for subsequent waves, we generated a large number of samples from the NROY space before sub-sampling these using a *maximin* design. For a consistent comparison, we ran 10 waves of history matching for each example in Section 5.

## 5 Results

To produce the model runs we are grateful to have been able to use JASMIN, the UK’s collaborative data analysis environment [Lawrence et al., 2013, https://jasmin.ac.uk]. We also acknowledge the Catalyst HPC cluster at the University of Bristol where we ran early tests of the model. Emulation and history matching were conducted on standard desktop/laptop machines.

Amongst all of the different scenarios below, the minimum time taken for a single particle filter to run was 1.7 hours, and the longest was 11.6 hours. To balance out HPC resource requests, we decided to run each of the 250 design/validation points in a given wave in parallel on JASMIN, over 250 independent nodes (so each design point was evaluated in serial). We note that given multiple cores on each node, runtimes could be further improved by parallelising various aspects of the particle filter, at the cost of requiring additional CPU capabilities. Nonetheless, evaluating the model, even with a small number of particles, is highly computationally expensive, and as such this further justifies the use of emulation methods.

### 5.1 Simulation study

As a test of the code we simulated an outbreak using a set of parameter inputs that were present in the initial design. In this case seeding infections were introduced randomly through the model discrepancy process in different spatial regions. We introduced a lockdown at day 37, and ran the simulation for 68 days in total. The parameters we used for the simulation are shown in Supplementary Table S1. We applied the observation process described in Section 3.4 to generate ‘observed’ data at the same levels of aggregation as seen in the real data. We then conducted 10 waves of history matching, using the approach described in Section 4.4, with 200 design points and 50 validation points at each wave (corresponding to a standard 80:20 split of training to validation samples). We only fitted the model to data up to day 54, but then simulated the particle trajectories forwards to generate probabilistic forecasts for the final two-weeks. The mean time to run the particle filter across all waves was 3.9 hours, with a range of 2.7–6.1 hours.

Figure 2 summarises the particle trajectories across the ensemble of design points at Wave 10 fitted to the simulated data at day 54. We can see that the epidemic started in Yorkshire and the North East, and that the particle trajectories seem to be tracking the observed data well. We note that the two-week forecasts also seem good here, with the uncertainty bounds containing the true (but unobserved) trajectories. For comparison, Supplementary Figure S1 shows the particle trajectories over the initial Wave 1 input space, and here we can see that even with the filtering the particle trajectories have a very large variance over the initial plausible region. This illustrates the improvement in model fit through successive waves of history matching. We reiterate that there is *no explicit seeding process* in this model, instead we simply allow the MD process to introduce infections in the early stages and then let the particle filter guide the trajectories using information in the available data. We note also that we can obtain estimates for all the underlying hidden states, which for the Wave 10 particles are shown in Supplementary Figure S6. We can see that these match the true (but hidden) trajectories fairly well. It is worth noting that the model is predicting fewer hospital deaths in the oldest age-category than the true trajectory, but since the model is only being fitted to total deaths (i.e. *D*_*I*_ + *D*_*H*_), there is limited information in the data to identify the true trajectories of *D*_*I*_ and *D*_*H*_ separately.

**Figure 2:**
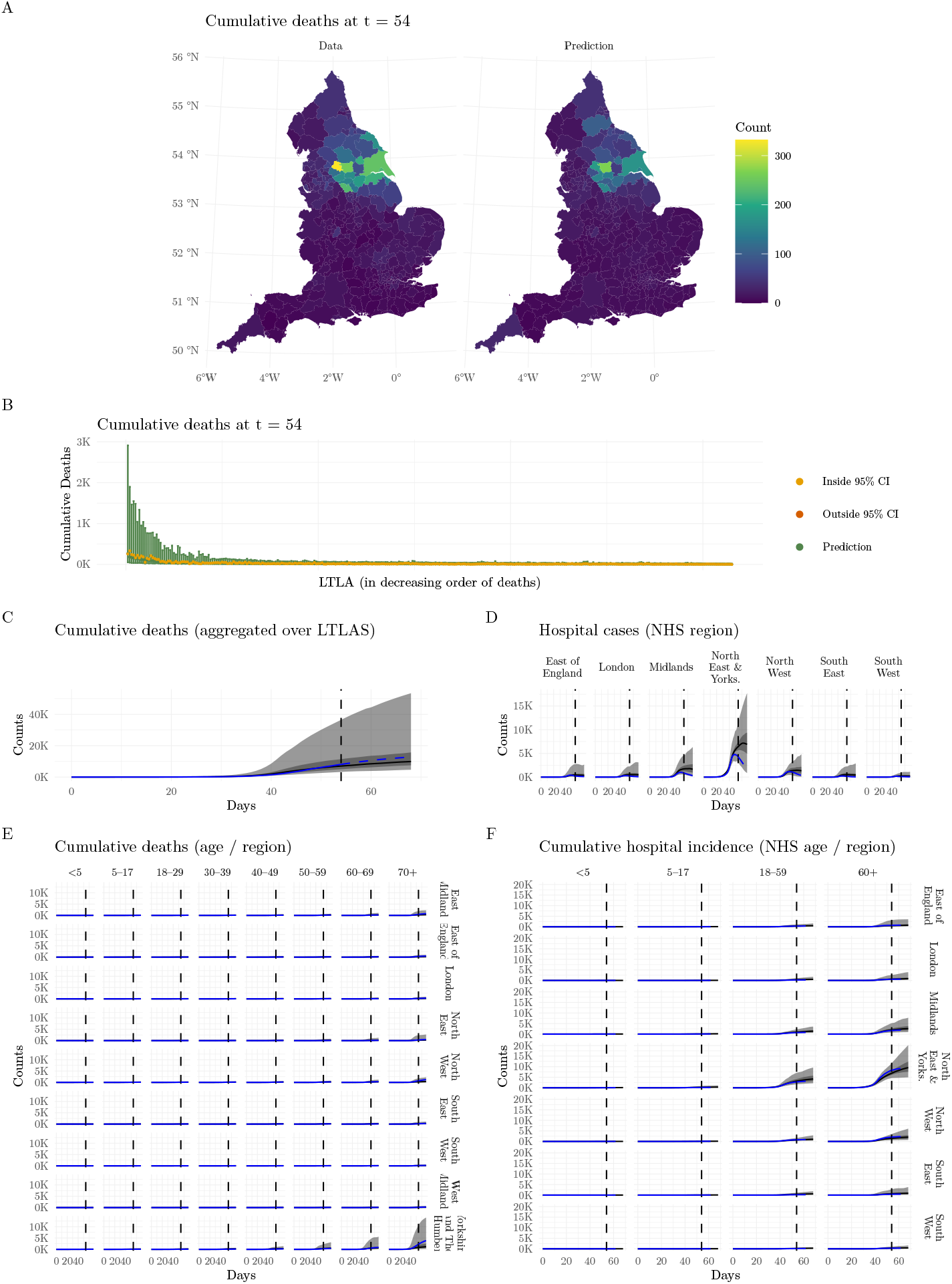
Particle trajectory plots across the ensemble of design points at Wave 10 for the simulated outbreak. A) Spatial (LTLA-level) plots of the mean number of deaths by day 54, for the data and the fitted model. B) Cumulative deaths at day 54 within each LTLA (ranked in decreasing order of predicted deaths). C) Cumulative deaths over time, aggregated over the 338 LTLAs. D) Hospital cases over time in each NHS region. E) Cumulative deaths over time in each age/region category. F) Cumulative hospital incidence over time by each NHS age/region category. In plot B) the green points are the predicted ensemble means, and the error bars are the 95% prediction intervals. The yellow and red points are the observed data coloured by whether they lie inside and outside of the prediction intervals respectively. In plots C–F, the blue dashed lines correspond to the observed data and the black solid lines to the mean trajectories from the particles taken across the ensemble. The ribbons correspond to 50% and 95% prediction intervals. A vertical dashed line corresponds to the end point of the observed data, such that trajectories before the line are generated from the particle filter, and trajectories to the right of the line are simulated forecasts from the model.

Supplementary Figures S10 and S11 show the evolution of the NROY space over the different waves for the parameters of the model, with the true values of the parameters shown as red points. These show that over successive waves the NROY space becomes a small proportion (≈ 1.7 × 10^−9^) of the original input space, and that at Wave 10 the true values are contained within the NROY space, suggesting that the HM process is honing in on the correct region of parameter space. We can also see that we cut more space out for some variables than others, and this is likely due to some variables remaining inactive across the different waves. We discuss this in more detail in Section 6. Overall we can see that the approach is working well.

#### 5.1.1 Simulation study assuming availability of data at the correct model aggregations

We also ran 10 waves of history matching on the simulated data assuming that the observed hospital and death data was available at the same resolution as the model (i.e. at the age / LTLA level). The mean time to run the particle filter across all waves was 4 hours, with a range of 2.9–11.6 hours. For brevity the results are shown in the Supplementary Materials, with the ensemble particle trajectories for Waves 1 and 10 shown in Figures S4 and S5 respectively. Again, by Wave 10 the model is doing a good job of fitting to all spatial locations in all age-classes, except perhaps the number of hospital deaths in the oldest age-class, where the model is underpredicting the number of deaths in the LTLAs with the highest observed number of deaths at day 54. In Section 5.1 we noted that the previous model underestimated the ‘true’ number of hospital deaths, most likely due to the model being fitted only to total observed deaths, and not by hospital and community deaths simultaneously (e.g. Figure S6). If we look at the predicted hidden states for this new model—Figure S9—we can see that the model is still underpredicting the *D*_*H*_ state, but not by as much as the previous model, suggesting (as expected) that this increase of information in the observed data results in better predictions of the hidden states, even if not perfect. To explore this behaviour in more detail, we can also see that once again at Wave 10 the NROY space is a small proportion (≈ 1.1 × 10^−8^) of the original input space (Figures S16–S17), but in particular the variables *ν*_*I*_ and *ν*_*H*_ (which allow the relative hospital and community death rates by age to vary) are not yet well-identified, even with the increased information in the data. The fits could potentially be improved by running more waves of history matching, however in this case only 12% of the NROY space was removed after Wave 9, and examination of the validation plots from the emulator suggested that the variance between log-likelihood estimates across the NROY space is high enough that we are unlikely to reduce space substantially more without reducing the Monte Carlo error of the particle filter estimates. We will return to this challenge in Section 6.

### 5.2 Real data

For the real data we have the additional complexity that the hospitalisation data is only available from 19^th^ March 2020, whereas the death data is available from 2^nd^ March 2020. Hence the likelihood terms for the observation process need to be adjusted to target only the available data at any given time point.

#### 5.2.1 Fitted to data up to the first lockdown

Since we are unsure of the initial conditions, we choose to run the model from 15^th^ February 2020, up to the first lockdown on the 23^rd^ March 2020, a total of 37 days. We assume that deaths from COVID-19 are zero between 15^th^ February–2^nd^ March, but leave the hospitalisation data missing until 19^th^ March. We assume the population was fully susceptible on the 15^th^ February, and allow the MD process to introduce infections as required. We then produce forecasts for the following two-weeks.

As before we conducted 10 waves of history matching, and the mean time to run the particle filter across all waves was 2.9 hours, with a range of 1.7–5.3 hours. The particle trajectories for Wave 10 are summarised in Figure 3, and a comparison to the Wave 1 trajectories (Supplementary Figure S2) shows a substantial improvement in model fit over successive waves of history matching. Here we can see that the model is doing a good job of estimating the spatial heterogeneity in the observed numbers of deaths, and is generally fitting most observed trajectories well. It seems to overpredict the cumulative deaths in various LTLAs (Figure 3B), resulting in a smoother distribution over space than seen in the data (Figure 3A). This could be for various reasons, the predominant one being that there is much less information in the dataset at this point than we had in the simulated data, especially since hospitalisations are only available 4 or 5 days prior to the first lockdown. This has the knock-on effect that in the early stages of the outbreak the only data that can be used to inform the particle weights is found right at the end of the epidemiological pathways described in Figure 1. Given a finite number of particles this can lead to a lag-effect, such that by the time there is sufficient information in the data to constrain the trajectories, they are already moving away from the truth. We note that this is a generic challenge with using boostrap particle filters, but is felt more acutely here due to the lack of hospitalisation data in the early stages, and the small number of particles that we are able to run. We discuss these challenges in more detail, and possible ways they could be alleviated in future work in Section 6.

**Figure 3:**
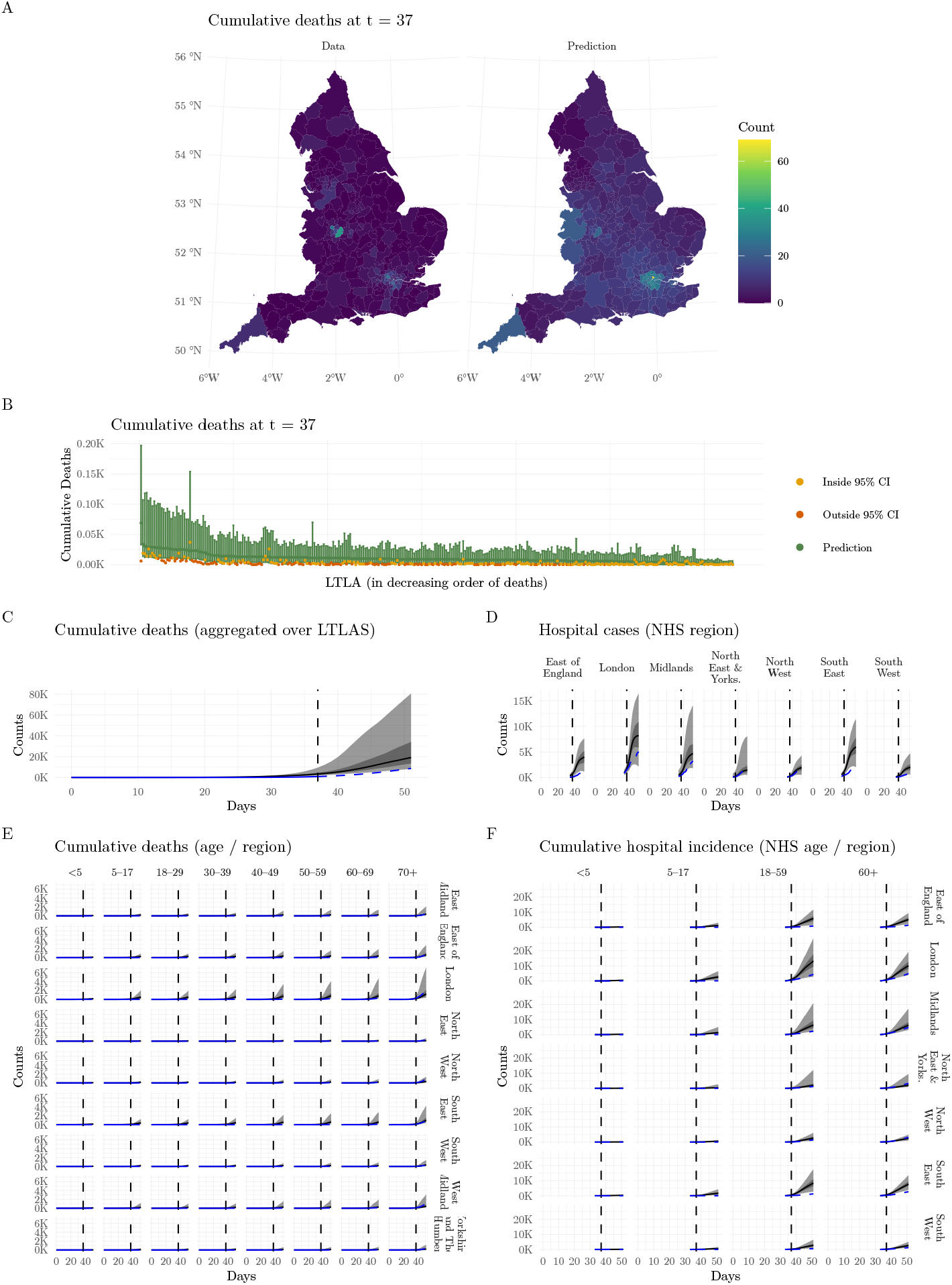
Particle trajectory plots across the ensemble of design points at Wave 10 for the real UK data up to the first lockdown. A) Spatial (LTLA-level) plots of the mean number of deaths by day 37, for the data and the fitted model. B) Cumulative deaths at day 37 within each LTLA (ranked in decreasing order of predicted deaths). C) Cumulative deaths over time, aggregated over the 338 LTLAs. D) Hospital cases over time in each NHS region. E) Cumulative deaths over time in each age/region category. F) Cumulative hospital incidence over time by each NHS age/region category. In plot B) the green points are the predicted ensemble means, and the error bars are the 95% prediction intervals. The yellow and red points are the observed data coloured by whether they lie inside and outside of the prediction intervals respectively. In plots C–F, the blue dashed lines correspond to the observed data and the black solid lines to the mean trajectories from the particles taken across the ensemble. The ribbons correspond to 50% and 95% prediction intervals. A vertical dashed line corresponds to the end point of the observed data, such that trajectories before the line are generated from the particle filter, and trajectories to the right of the line are simulated forecasts from the model.

Supplementary Figure S12 shows the input spaces for the *t*_*r*,MD_ terms, and we can see very clearly the spatial variation, with London requiring MD to be added earlier than the rest of the country, and the North West much later. Supplementary Figure S13 shows the rest of the parameters, and we can see that by Wave 10 the NROY space is a very small proportion (≈ 1.4 *×* 10^−13^) of the original input space, despite the paucity of available data compared to the simulation study.

#### 5.2.2 Fitted to data beyond the first lockdown

After the first lockdown comes into place on 23^rd^ March 2020, we introduce additional parameters to capture the impacts of control measures: firstly, work and play movements happen at random with a probability *p*_move_; and secondly, the force-of-infection in (1) is multiplied by a parameter 0 *< β*_*s*_ *<* 1, to model the impact of interventions such as social distancing. We also swap the contact matrices in (1) from one derived from the POLYMOD survey Mossong et al. [2008], to one derived from the CoMix survey after the 23^rd^ March 2020 Jarvis et al. [2020]. As before, we start the modelling on 15^th^ February and fit to data up to 9^th^ April 2020, corresponding to 54 days in total, with forecasts generated for a further two-weeks.

As before we conducted 10 waves of history matching, and the mean time to run the particle filter across all waves was 4.7 hours, with a range of 3–8.7 hours. The particle trajectories for Wave 10 can be seen in Figure 4, and a comparison to the Wave 1 trajectories (Supplementary Figure S3) again shows a substantial improvement in model fit over successive waves of history matching. Here we can see that the information afforded by the additional 17 days’ worth of data has substantially improved the model fits. In Figure 4B we can again see the lag-effect that was discussed in the previous section, where all the trajectories seem to overpredict in the early stages but then pull back to align with the observed data later on. This is further evidence that we need better particle filters, since it may be that we are inducing some bias in the parameter inference as a means of correcting for the early lag-effect. Supplementary Figures S14 and S15 show the evolution of the NROY space over successive waves, and we can again see that the Wave 10 NROY space is a very small proportion (≈ 5.3 *×* 10^−14^) of the original input space.

**Figure 4:**
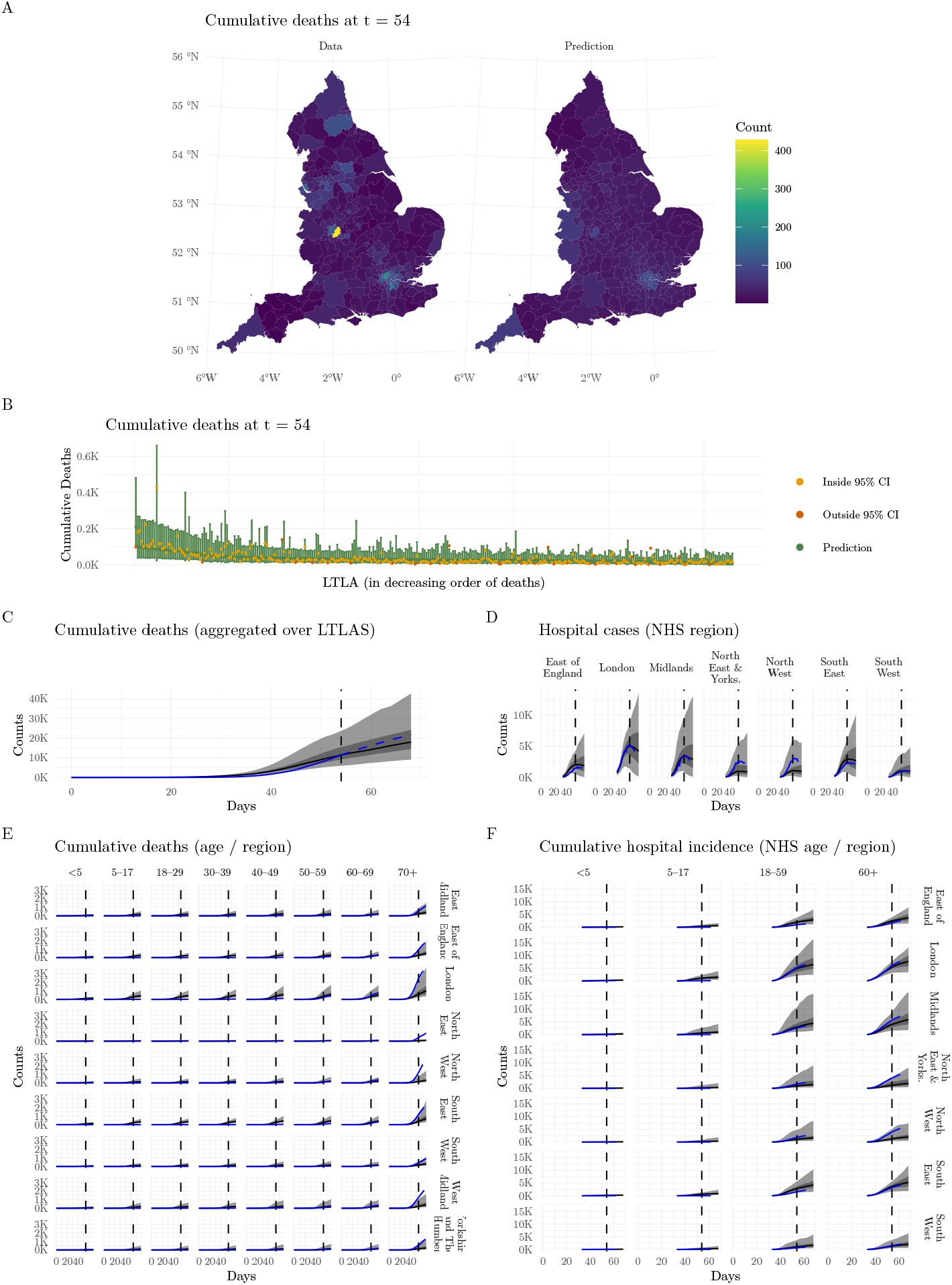
Particle trajectory plots across the ensemble of design points at Wave 10 for the real UK data beyond the first lockdown. A) Spatial (LTLA-level) plots of the mean number of deaths by day 54, for the data and the fitted model. B) Cumulative deaths at day 54 within each LTLA (ranked in decreasing order of predicted deaths). C) Cumulative deaths over time, aggregated over the 338 LTLAs. D) Hospital cases over time in each NHS region. E) Cumulative deaths over time in each age/region category. F) Cumulative hospital incidence over time by each NHS age/region category. In plot B) the green points are the predicted ensemble means, and the error bars are the 95% prediction intervals. The yellow and red points are the observed data coloured by whether they lie inside and outside of the prediction intervals respectively. In plots C–F, the blue dashed lines correspond to the observed data and the black solid lines to the mean trajectories from the particles taken across the ensemble. The ribbons correspond to 50% and 95% prediction intervals. A vertical dashed line corresponds to the end point of the observed data, such that trajectories before the line are generated from the particle filter, and trajectories to the right of the line are simulated forecasts from the model.

## 6 Discussion

We have developed a novel method for calibrating computationally intensive stochastic infectious disease models in high-dimensions, that utilises emulation and history matching to perform robust exploration of the parameter space to find the region of high posterior density. Using this approach we have successfully calibrated an age-structured model (8 age-classes) across a meta-population of 338 connected spatial regions, where the underlying simulation model also includes patterns of movement driven by commuter data and a transmission process governed by contact matrices derived from social mixing surveys. Traditional approaches to this problem require emulators to be built for each output-of-interest, which would be prohibitive here since we want to fit to time-series data across 338 spatial areas and multiple time points simultaneously. Alternatively one could fit to aggregated data [e.g. Vernon et al., 2022], but this would lose the ability to accurately track epidemic trajectories in high-dimensions. We circumvent this problem by emulating a transformation of the log-likelihood surface instead, which is a one-dimensional output and thus only requires a single emulator [see also Oakley and Youngman, 2017, Wilkinson, 2014].

However, as with almost all infectious disease systems the likelihood function is analytically intractable, since it’s evaluation relies on inferring a (often large) set of hidden states [e.g. Gibson and Renshaw, 1998, O’Neill and Roberts, 1999, Jewell et al., 2009, Deardon et al., 2010]. We approach this problem by using particle filtering to generate estimates of the likelihood that rely only on being able to simulate directly from the underlying complex model [Gordon et al., 1993]. Furthermore we develop a novel model structure where we embed a model discrepancy process into the simulation pipeline, which, alongside the use of particle filtering tackles various challenges, particularly how to deal with introductions of infections from processes that are not explicitly captured in the simulation model (including imports from outside of the UK, which is vital in the early stages of the outbreak to generate plausible seeding in time and space and for which a suitable mechanistic model does not exist).

Particle filtering (and other simulation-based) methods have been used successfully in the past for calibrating infectious disease models, usually by embedding within MCMC [see e.g. Andrieu and Roberts, 2009, Andrieu et al., 2010, McKinley et al., 2014, Del Moral et al., 2015, Drovandi et al., 2016, McKinley et al., 2020] or SMC^2^ [Chopin et al., 2012] algorithms. However, these methods can be extremely computationally challenging, often requiring large numbers of particles to be run, even in relatively small-scale systems. In the examples explored here, the minimum runtime for a single particle filter evaluation was 1.7 hours, simulating across a 37 day period, but the longest was 11.6 hours simulating across a 54 day period. In future work we would aim to fit models to even longer time periods, and all of this together make it infeasible to embed these kinds of filters in standard MCMC or SMC algorithms.

As a result, we instead use emulation as a fast way to comprehensively explore the input space, and combine this with iterative history matching to efficiently converge towards the area of high posterior density through a series of waves (since under the uniform priors used here the posterior density is proportional to the likelihood). An emulator can be trained from a relatively small set of training runs of the model, and then used to systematically explore the parameter space to remove areas of the input space where the probability of the true likelihood being within a specified region around the posterior mode is low (the implausible region). These training runs are can be easily parallelised across design points, and/or within the particle filters as required. The number of training points can be fixed in advance, meaning that the computational burden can be controlled more effectively on HPC clusters. Once trained, the emulators are quick to evaluate, and so large numbers of predictions can be made quickly and efficiently, meaning that the emulator can comprehensively explore the input space in a way that can’t be achieved if we were to rely on the particle filters alone. Log-likelihood surfaces can be challenging to emulate, so we exploit recent advances for efficiently and robustly fitting Deep Gaussian Processes to do this [Ming et al., 2023].

We fit to publicly available, but incomplete hospitalisation and death data that are available at different degrees of spatial/age-structured hierarchies. Overall we used only 2,500 evauations of the particle filters for each scenario we explored, with the emulators being built and run on standard machines. Although not perfect, the fitted models show good calibration in space-and-time, although with key areas for improvement in future work, as discussed in detail below.

A key novelty we introduce is to embed a structured model discrepancy process into the system. We set this up as an idealised zero-mean noise process that operates across all epidemiological states and allows a mechanism for amending the hidden states based on processes that are not explicitly modelled, such as the introduction of infection in the early stages of the outbreak. *This latter consideration was a major challenge to overcome*. Running the full model in the prior space produced some runs that were consistent with the data at the national level, but not at finer spatial resolutions (not even at the regional-level let alone the LTLA-level). One way to approach this issue is to introduce some spatio-temporal seeding process, such as to have some spatially-explicit background rate-of-infection parameters that exert a constant infectious pressure over time. There are various problems with this approach: firstly, there is little evidence to inform the functional form of these terms (there certainly isn’t a constant pressure in reality), and secondly, this introduces a large number of additional parameters to estimate which increases the volume of the input space dramatically. We tried a range of other approaches for generating seeding events heterogeneously across spatial regions, including a method based on using the first few weeks worth of data to generate empirical priors for the likely states of the system at a fixed point in time by running simpler MCMC models in each spatial region independently. None of approaches we tried worked well, but the embedded MD process, coupled with the use of particle filtering, provided a flexible approach to dealing with this problem, and we have shown in this paper that the method can produce good model fits even without any explicit seeding mechanism in the model. This differs from other approaches to dealing with unknown mechanisms because it allows for model states to be increased *or* decreased with respect to information in the observed data. It also ensures that the added noise results in epidemiologically consistent hidden states. Furthermore, the use of MD also introduces additional uncertainty that represents the fact that the underlying transmission model structure is itself an approximation of reality. The ability to track trajectories in space and time is of vital importance if we want to use models to produce spatio-temporal predictions/forecasts, and/or to explore control policies such as localised lockdowns.

However, there are still key challenges to overcome. The main one relates to the effectiveness of the particle filter to build in information across the whole time-series when simulating particles. For example, in Section 3.3.2 we discuss the introduction of a set of region-specific parameters controlling the amount of MD error introduced in the early stages of the outbreak. We found that without these parameters the particle trajectories would systematically overestimate the observed curves, despite, in theory, the MD process being able to increase or decrease the numbers of individuals in each state over time. So, what is happening here? The first thing to note is that although the MD has an idealised zero-mean, in order to ensure that the MD adjusted states are epidemiologically valid we require truncation of the MD distributions (see Section 3.3). When counts are low, and truncated below by zero, this necessarily means that the mean of the truncated MD process is *>* 0, which makes epidemiological sense, since the counts can’t be less than zero. However, it also means that in the early stages there is a higher likelihood of the MD increasing the counts rather than decreasing them (i.e. the necessary truncation induces some skewness). As such, in the initial stages particles are more likely to increase on average. The second thing to note is that in the early stages of the outbreak there are negligible control measures in place, and in that situation *R*_0_ is larger than 1 (in fact *R*_0_ *>* 2 in our model). This means that once seeds are introduced the simulation model will kick in and drive infections upwards unless there is information in the data to put preferential weight on lower counts. This leads to the third consideration, which is that the observed data relate to epidemiological states found towards the *ends* of the various pathways described in Figure 1 (i.e. hospitalisations and deaths). This induces a *lag-effect*, which the standard BPF cannot overcome since it has no way of adjusting states at earlier time points based on data at much later time points. This is a well-known problem in particle filtering, but is acutely felt here because we are trying to track trajectories in high dimensions, in a highly stochastic model with a lag-effect induced by the observable data. We note that this issue is a practical consideration that would be unlikely to happen if we were able to run very large numbers of particles, since there would be a non-negligible probability that at least some of the particles were on trajectories more consistent with later time points.

We deal with this issue in part by introducing the parameters described in Section 3.3.2, which slows the introduction of MD in different regions. In practice this controlled things better at the regional level (as seen in the various posterior predictive plots such as Figures 2–4), but it is likely that some of these issues were still felt at the LTLA-level, where individual LTLAs were not always mapped precisely, despite the regional-level predictions being good. This could of course be alleviated by having a finer spatial resolution for when the MD process begins to kick in, but this comes at the cost of increasing the size of the parameter space, which is one of the key things we wish to avoid having to do.

All of this motivates future work to explore the use of more sophisticated look-ahead particle filters. A key consideration is that although it is straightforward to simulate forwards from the transmission model, it is hard to condition those simulations on observed data without completely rewriting the code base (this is especially challenging given e.g. stochastic movements of individuals between regions). In simulation work we were able to design a one-step ahead auxiliary particle filter [Pitt and Shephard, 1999] that helped alleviate this problem a bit by simulating the ***X***_*t*_ terms directly from the transmission model, and then using the observed data ***Z***_*t*_ to inform the simulation of the MD adjusted states, ***Y***_*t*_, given ***X***_*t*_ and ***Z***_*t*_. Since the ***Y***_*t*_ | ***X***_*t*_ are independent discrete Gaussian random variables, it is easier to build information from ***Z***_*t*_ into the simulation of ***Y***_*t*_ | ***X***_*t*_ than it is to build it into the simulation of ***X***_*t*_ | ***Y***_*t*_ as would be the usual approach. However, it was challenging to get this working for the real data due to the fact that the observed data were also specified on a different scale to the hidden states (since the one-step ahead filters use the future data to inform simulation of earlier hidden states, this requires a way to map information in observed data at one level of aggregation down to the hidden states at another level of aggregation, which is highly challenging for a model of this complexity). As such, we didn’t progress much further with this here.

Other recent advances in the field include twisted particle filters (TPFs; Guarniero et al., 2017), which are designed explicitly to deal with this exact issue, where information at later time points can be fed back to inform simulations at earlier time points through the introduction of twisting functions that amend the filtering distributions accordingly. These approaches maintain the unbiasedness properties of the likelihood estimate but can reduce the Monte Carlo variance of these estimates hugely. The cost is that the twisting functions have to be learned, and so approaches such as that of Guarniero et al. [2017] iterate through each particle filter multiple times in order to learn these twisting functions appropriately. For a system such as ours, where a single evaluation of a particle filter is highly time consuming, then this could be computationally prohibitive. However, using emulation and history matching helps to alleviate some of these computational challenges by reducing the number of filters that have to be evaluated, certainly compared to the alternative of embedding a TPF into an MCMC or SMC^2^ algorithm. By reducing the variance in the likelihood estimates it may be possible to cut out substantially more space at each wave of history matching, thus requiring less training runs of the model to calibrate effectively. We also think that it is likely that with a good look-ahead filter it would not be necessary to include additional parameters controlling when MD is added, such as those developed in Section 3.3.2, thus making the calibration challenge far easier.

This is a key area of interest for future research, because an efficient look-ahead filter, coupled with the embedded MD process we have developed, would provide a powerful way to deal with many practical issues associated with calibrating complex spatio-temporal infectious disease models that we have discussed in this paper. We note that key challenges would also have to be overcome in terms of mapping the observed data at low spatial resolutions to the hidden states at high spatial resolutions, as previously discussed for the auxiliary particle filter. Additional methodological work would be required to figure out how to twist the complex transmission model, which is defined in discrete-time and space, and further compounded by the additional MD process. It may be possible to employ similar ideas to those discussed above for the APF, or leverage ideas around conditional sampling similar to those developed in Supplementary Materials S4. Another interesting approach would be to explore whether it would be possible to target an approximate likelihood surface that can be evaluated using more tractable (but approximate) models [e.g. Whiteley and Rimella, 2021]. Another area-of-interest would be how to perform *online* inference, since the current approach requires simulating the complete time-series each time one wishes to update the calibration with new data, which is likely to become very time-consuming for long outbreaks.

Overall we think that the broad approach developed here and in Part 1 [Williamson et al., 2025] show great potential in terms of being a generalisable way to calibrate complex infectious disease models, and if efficient look-ahead filtering approaches could be developed then this may well move this into the realms of being able to perform robust inference in real-time.

## Data Availability

All data produced in the present study are available upon reasonable request to the authors.

