## Supplementary Materials for "On real-time calibrated prediction for complex model-based decision support in pandemics: Part 2"

#### S1 Packages used

R (R Core Team, 2022) packages used in this work include: `mclust` (Scrucca et al., 2023), `tidyverse` (Wickham et al., 2019), `lubridate` (Grolemund and Wickham, 2011), `GGally` (Schloerke et al., 2024), `truncnorm` (Mersmann et al., 2023), `patchwork` (Pedersen, 2022), `Rcpp` (Eddelbuettel and François, 2011), `RcppArmadillo` (Eddelbuettel and Sanderson, 2014), `abind` (Plate and Heiberger, 2024), `sitmo` (Balamuta et al., 2021), `sf` (Pebesma, 2018), `lhs` (Carnell, 2024), `areal` (Prener and Revord, 2019), `hmer` (Iskauskas et al., 2024), `MASS` (Venables and Ripley, 2002), `fields` (Nychka et al., 2021), `dgpsi` (Ming and Williamson, 2024), `data.table` (Barrett et al., 2025), `NIMBLE` (de Valpine et al., 2017) and `R.utils` (Bengtsson, 2025).

#### S2 Mapping $R_0$ to $\nu$ using the next-generation matrix (NGM)

We refer the reader to e.g. Diekmann et al. (1990); van den Driessche and Watmough (2002); Diekmann et al. (2010); van den Driessche (2017) for more details of the method of constructing NGMs for compartmental models, and using them to derive equations for $R_0$. Here we will use the construction of van den Driessche (2017).

Firstly, let $F_i(x)$ be the rate of *new* infections into *infected* class $i$, and $V_i(x)$ be the rate of transitions from *infected* class $i$. Then define matrices:

$$\mathbf{F} = \left[\frac{\partial F_i\left(x_0\right)}{\partial x_j}\right] \quad \text{and} \quad \mathbf{V} = \left[\frac{\partial V_i\left(x_0\right)}{\partial x_j}\right] \quad \text{for } 1 \leq i, j, \leq N_{\text{inf}},$$

where $x_0$ corresponds to the states at the disease-free equilibrium and $N_{\text{inf}}$ is the number of infected classes. Then, the NGM, $\mathbf{K}$, is:

$$\mathbf{K} = \mathbf{F}\mathbf{V}^{-1},$$

and $R_0$ is the maximum eigenvalue of $\mathbf{K}$.

From the model structure above, in a single population with age-structure, the **infected classes** are $X_{a,E}$, $X_{a,A}$, $X_{a,P}$, $X_{a,I_1}$ and $X_{a,I_2}$ for age-classes $a = 1, \ldots, N_a$. Note that from a *biological* perspective individuals in hospital are also infected, but in our model they play no further role in the transmission process, and so we do not count $X_{a,H}$ as an infected state for the purposes of constructing the NGM. In a *deterministic* model the infected states above would have rates-of-change of:

$$\frac{dX_{a,E}}{dt} = X_{a,S}\left[\sum_{k=1}^{N_a}\frac{\beta_{ka}}{M_k}\left(X_{k,P}+X_{k,I_1}+X_{k,I_2}+\nu_A X_{k,A}\right)\right]-\gamma_E X_{a,E}$$
$$\frac{dX_{a,A}}{dt} = \left(1-p_{a,EP}\right)\gamma_E X_{a,E}-\gamma_A X_{a,A}$$
$$\frac{dX_{a,P}}{dt} = p_{a,EP}\gamma_E X_{a,E}-\gamma_P X_{a,P}$$
$$\frac{dX_{a,I_1}}{dt} = \gamma_P X_{a,P}-\gamma_{I_1} X_{a,I_1}$$
$$\frac{dX_{a,I_2}}{dt} = \left(1-p_{a,I_1H}-p_{a,I_1D}\right)\gamma_{I_1} X_{a,I_1}-\gamma_{I_2} X_{a,I_2}$$

Here $\beta_{ka}$ is the transmission rate from class $k$ to class $a$ and $M_k$ is the total number of individuals in class $k$. The parameter $\nu_A \in (0,1)$ scales the force-of-infection from the $A$ class relative to the other infectious classes.

Letting $i = 1,\dots,N_a$ and $j = 1,\dots,N_a$, then for brevity we only show the non-zero components needed to derive the NGM, which are:

$$\frac{\partial F_{X_{i,E}}\left(x_0\right)}{\partial X_{j,A}} = \frac{\beta_{ji}\nu_A X_{i,S}}{M_j} \qquad \forall i,j$$
$$\frac{\partial F_{X_{i,E}}\left(x_0\right)}{\partial X_{j,P}} = \frac{\beta_{ji} X_{i,S}}{M_j} \qquad \forall i,j$$
$$\frac{\partial F_{X_{i,E}}\left(x_0\right)}{\partial X_{j,I_1}} = \frac{\beta_{ji} X_{i,S}}{M_j} \qquad \forall i,j$$
$$\frac{\partial F_{X_{i,E}}\left(x_0\right)}{\partial X_{j,I_2}} = \frac{\beta_{ji} X_{i,S}}{M_j} \qquad \forall i,j$$

and

$$\frac{\partial V_{X_{i,E}}\left(x_0\right)}{\partial X_{j,E}} = \begin{cases} \gamma_E & \text{for } i=j, \\ 0 & \text{otherwise,} \end{cases}$$
$$\frac{\partial V_{X_{i,A}}\left(x_0\right)}{\partial X_{j,E}} = \begin{cases} -\left(1-p_{i,EP}\right)\gamma_E & \text{for } i=j, \\ 0 & \text{otherwise,} \end{cases}$$
$$\frac{\partial V_{X_{i,A}}\left(x_0\right)}{\partial X_{j,A}} = \begin{cases} \gamma_A & \text{for } i=j, \\ 0 & \text{otherwise,} \end{cases}$$
$$\frac{\partial V_{X_{i,P}}\left(x_0\right)}{\partial X_{j,E}} = \begin{cases} -p_{i,EP}\gamma_E & \text{for } i=j, \\ 0 & \text{otherwise,} \end{cases}$$
$$\frac{\partial V_{X_{i,P}}\left(x_0\right)}{\partial X_{j,P}} = \begin{cases} \gamma_P & \text{for } i=j, \\ 0 & \text{otherwise,} \end{cases}$$
$$\frac{\partial V_{X_{i,I_1}}\left(x_0\right)}{\partial X_{j,P}} = \begin{cases} -\gamma_P & \text{for } i=j, \\ 0 & \text{otherwise,} \end{cases}$$
$$\frac{\partial V_{X_{i,I_1}}\left(x_0\right)}{\partial X_{j,I_1}} = \begin{cases} \gamma_{I_1} & \text{for } i=j, \\ 0 & \text{otherwise,} \end{cases}$$
$$\frac{\partial V_{X_{i,I_2}}\left(x_0\right)}{\partial X_{j,I_1}} = \begin{cases} -\left(1-p_{i,I_1H}-p_{i,I_1D}\right)\gamma_{I_1} & \text{for } i=j, \\ 0 & \text{otherwise,} \end{cases}$$
$$\frac{\partial V_{X_{i,I_2}}\left(x_0\right)}{\partial X_{j,I_2}} = \begin{cases} \gamma_{I_2} & \text{for } i=j, \\ 0 & \text{otherwise.} \end{cases}$$

Since the non-zero components of $\mathbf{F}$ all contain $\beta_{ji}$, where

$$\beta_{ji} = \nu c_{ji},$$

we can therefore write

$$\mathbf{K} = \nu\mathbf{G}\mathbf{V}^{-1}$$

where $\mathbf{G}$ is equivalent to replacing $\beta_{ji}$ by $c_{ji}$ in $\mathbf{F}$. As such:

$$R_0 = \nu \ \text{eig}_M \left(\mathbf{G}\mathbf{V}^{-1}\right)$$
$$\Rightarrow \nu = \frac{R_0}{\text{eig}_M \left(\mathbf{G}\mathbf{V}^{-1}\right)}$$

where $\text{eig}_M (\mathbf{K})$ denotes the maximum eigenvalue of $\mathbf{K}$.

When using a mixture of contact matrices, we parameterise $\nu$ using the NGM evaluated at the initial contact structure, which represents the contact matrix expected in a completely susceptible population at the start of the outbreak.

### S3 Model discrepancy constraints and updates

Starting with the absorbing states we have:

$$\begin{aligned}
\left(\Delta'_{tas,D_H} \mid \boldsymbol{X}_{tas}, \boldsymbol{Y}_{(t-1)as}\right) &\sim N^D\left(0, \sigma^2_{tas,D_H}\right) I\left(-X'_{tas,D_H}, Y_{(t-1)as,H} - X'_{tas,D_H}\right),\\
\sigma^2_{tas,D_H} &= 2a_{\text{MD}} + 2b_{\text{MD}} X'_{tas,D_H},\\
Y'_{tas,D_H} &= X'_{tas,D_H} + \Delta'_{tas,D_H},\\
Y_{tas,D_H} &= X_{tas,D_H} + \Delta'_{tas,D_H}.
\end{aligned} \tag{S.1}$$

$$\begin{aligned}
\left(\Delta'_{tas,D_I} \mid \boldsymbol{X}_{tas}, \boldsymbol{Y}_{(t-1)as}\right) &\sim N^D\left(0, \sigma^2_{tas,D_I}\right) I\left(-X'_{tas,D_I}, Y_{(t-1)as,I_1} - X'_{tas,D_I}\right),\\
\sigma^2_{tas,D_I} &= 2a_{\text{MD}} + 2b_{\text{MD}} X'_{tas,D_I},\\
Y'_{tas,D_I} &= X'_{tas,D_I} + \Delta'_{tas,D_I},\\
Y_{tas,D_I} &= X_{tas,D_I} + \Delta'_{tas,D_I}.
\end{aligned} \tag{S.2}$$

$$\begin{aligned}
\left(\Delta'_{tas,R_H} \mid Y'_{tas,D_H}, \boldsymbol{X}_{tas}, \boldsymbol{Y}_{(t-1)as}\right) &\sim N^D\left(0, \sigma^2_{tas,R_H}\right) I\left(-X'_{tas,R_H}, Y_{(t-1)as,H} - Y'_{tas,D_H} - X'_{tas,R_H}\right),\\
\sigma^2_{tas,R_H} &= 2a_{\text{MD}} + 2b_{\text{MD}} X'_{tas,R_H},\\
Y'_{tas,R_H} &= X'_{tas,R_H} + \Delta'_{tas,R_H},\\
Y_{tas,R_H} &= X_{tas,R_H} + \Delta'_{tas,R_H}.
\end{aligned} \tag{S.3}$$

$$\begin{aligned}
\left(\Delta'_{tas,R_I} \mid \boldsymbol{X}_{tas}, \boldsymbol{Y}_{(t-1)as}\right) &\sim N^D\left(0, \sigma^2_{tas,R_I}\right) I\left(-X'_{tas,R_I}, Y_{(t-1)as,I_2} - X'_{tas,R_I}\right),\\
\sigma^2_{tas,R_I} &= 2a_{\text{MD}} + 2b_{\text{MD}} X'_{tas,R_I},\\
Y'_{tas,R_I} &= X'_{tas,R_I} + \Delta'_{tas,R_I},\\
Y_{tas,R_I} &= X_{tas,R_I} + \Delta'_{tas,R_I}.
\end{aligned} \tag{S.4}$$

$$\begin{aligned}
\left(\Delta'_{tas,R_A} \mid \boldsymbol{X}_{tas}, \boldsymbol{Y}_{(t-1)as}\right) &\sim N^D\left(0, \sigma^2_{tas,R_A}\right) I\left(-X'_{tas,R_A}, Y_{(t-1)as,A} - X'_{tas,R_A}\right),\\
\sigma^2_{tas,R_A} &= 2a_{\text{MD}} + 2b_{\text{MD}} X'_{tas,R_A},\\
Y'_{tas,R_A} &= X'_{tas,R_A} + \Delta'_{tas,R_A},\\
Y_{tas,R_A} &= X_{tas,R_A} + \Delta'_{tas,R_A}.
\end{aligned} \tag{S.5}$$

Now moving on to the non-absorbing states, we have:

$$\begin{aligned}
\left(\Delta_{tas,H} \mid, Y_{tas,D_H}, Y_{tas,R_H}, \boldsymbol{X}_{tas}, \boldsymbol{Y}_{(t-1)as}\right) &\sim N^D\left(0, \sigma^2_{tas,H}\right)\\
&\quad I\left(-X_{tas,H} + Y_{(t-1)as,H} - Y'_{tas,D_H} - Y'_{tas,R_H},\right.\\
&\qquad Y_{(t-1)as,I_1} - Y'_{tas,D_I} - X_{tas,H} + Y_{(t-1)as,H}\\
&\qquad \left. -Y'_{tas,D_H} - Y'_{tas,R_H}\right),\\
\sigma^2_{tas,H} &= 2a_{\text{MD}} + 2b_{\text{MD}} X_{tas,H},\\
Y_{tas,H} &= X_{tas,H} + \Delta_{tas,H},\\
Y'_{tas,H} &= X_{tas,H} + \Delta_{tas,H} - Y_{(t-1)as,H} + Y'_{tas,D_H} + Y'_{tas,R_H}.
\end{aligned} \tag{S.6}$$

$$
\begin{aligned}
\left(\Delta_{tas,I_2} \mid Y'_{tas,H}, Y'_{tas,D_I}, Y'_{tas,R_I}, \boldsymbol{X}_{tas}, \boldsymbol{Y}_{(t-1)as}\right) \sim N^D\left(0, \sigma^2_{tas,I_2}\right)& \\
I\left(-X_{tas,I_2} + Y_{(t-1)as,I_2} - Y'_{tas,R_I},\right.& \\
Y_{(t-1)as,I_1} - Y'_{tas,D_I} - Y'_{tas,H}& \\
\left.-X_{tas,I_2} + Y_{(t-1)as,I_2} - Y'_{tas,R_I}\right),& \\
\sigma^2_{tas,I_2} = 2a_{\mathrm{MD}} + 2b_{\mathrm{MD}}X_{tas,I_2},& \\
Y_{tas,I_2} = X_{tas,I_2} + \Delta_{tas,I_2},& \\
Y'_{tas,I_2} = X_{tas,I_2} + \Delta_{tas,I_2} - Y_{(t-1)as,I_2} + Y'_{tas,R_I}.&
\end{aligned} \tag{S.7}
$$

$$
\begin{aligned}
\left(\Delta_{tas,I_1} \mid Y'_{tas,I_2}, Y'_{tas,D_I}, Y'_{tas,H}, \boldsymbol{X}_{tas}, \boldsymbol{Y}_{(t-1)as}\right) \sim N^D\left(0, \sigma^2_{tas,I_1}\right)& \\
I\left(-X_{tas,I_1} + Y_{(t-1)as,I_1}\right.& \\
- Y'_{tas,I_2} - Y'_{tas,D_I} - Y'_{tas,H},& \\
Y_{(t-1)as,P} - X_{tas,I_1} + Y_{(t-1)as,I_1} - Y'_{tas,I_2}& \\
\left.-Y'_{tas,D_I} - Y'_{tas,H}\right),& \\
\sigma^2_{tas,I_1} = 2a_{\mathrm{MD}} + 2b_{\mathrm{MD}}X_{tas,I_1},& \\
Y_{tas,I_1} = X_{tas,I_1} + \Delta_{tas,I_1},& \\
Y'_{tas,I_1} = X_{tas,I_1} + \Delta_{tas,I_1} - Y_{(t-1)as,I_1} + Y'_{tas,I_2}& \\
+ Y'_{tas,D_I} + Y'_{tas,H}.&
\end{aligned} \tag{S.8}
$$

$$
\begin{aligned}
\left(\Delta_{tas,P} \mid Y'_{tas,I_1}, \boldsymbol{X}_{tas}, \boldsymbol{Y}_{(t-1)as}\right) \sim N^D\left(0, \sigma^2_{tas,P}\right)& \\
I\left(-X_{tas,P} + Y_{(t-1)as,P} - Y'_{tas,I_1},\right.& \\
\left.Y_{(t-1)as,E} - X_{tas,P} + Y_{(t-1)as,P} - Y'_{tas,I_1}\right),& \\
\sigma^2_{tas,P} = 2a_{\mathrm{MD}} + 2b_{\mathrm{MD}}X_{tas,P},& \\
Y_{tas,P} = X_{tas,P} + \Delta_{tas,P},& \\
Y'_{tas,P} = X_{tas,P} + \Delta_{tas,P} - Y_{(t-1)as,P} + Y'_{tas,I_1}.&
\end{aligned} \tag{S.9}
$$

$$
\begin{aligned}
\left(\Delta_{tas,A} \mid Y'_{tas,R_A}, Y'_{tas,P}, \boldsymbol{X}_{tas}, \boldsymbol{Y}_{(t-1)as}\right) \sim N^D\left(0, \sigma^2_{tas,A}\right)& \\
I\left(-X_{tas,A} + Y_{(t-1)as,A} - Y'_{tas,R_A},\right.& \\
Y_{(t-1)as,E} - Y'_{tas,P} - X_{tas,A}& \\
\left.+Y_{(t-1)as,A} - Y'_{tas,R_A}\right),& \\
\sigma^2_{tas,A} = 2a_{\mathrm{MD}} + 2b_{\mathrm{MD}}X_{tas,A},& \\
Y_{tas,A} = X_{tas,A} + \Delta_{tas,A},& \\
Y'_{tas,A} = X_{tas,A} + \Delta_{tas,A} - Y_{(t-1)as,A} + Y'_{tas,R_A}.&
\end{aligned} \tag{S.10}
$$

$$
\begin{aligned}
\left(\Delta_{tas,E} \mid Y'_{tas,P}, Y'_{tas,A}, \boldsymbol{X}_{tas}, \boldsymbol{Y}_{(t-1)as}\right) \sim N^D\left(0, \sigma^2_{tas,E}\right)& \\
I\left(-X_{tas,E} + Y_{(t-1)as,E} - Y'_{tas,P} - Y'_{tas,A},\right.& \\
\left.Y_{(t-1)as,S} - X_{tas,E} + Y_{(t-1)as,E} - Y'_{tas,P} - Y'_{tas,A}\right),& \\
\sigma^2_{tas,E} = 2a_{\mathrm{MD}} + 2b_{\mathrm{MD}}X_{tas,E},& \\
Y_{tas,E} = X_{tas,E} + \Delta_{tas,E},& \\
Y'_{tas,E} = X_{tas,E} + \Delta_{tas,E} - Y_{(t-1)as,E} + Y'_{tas,P} + Y'_{tas,A}& \\
Y_{tas,S} = Y_{(t-1)as,S} - Y'_{tas,E}.&
\end{aligned} \tag{S.11}
$$

Since the model discrepancy terms are assumed conditionally independent within each age-class and region, for brevity we drop the $as$ subscripts in the derivations below. Note that in all derivations we leverage the fact that all counts must be $\geq 0$. The parameters of each MD distribution are given in Section 3.3 in Part 2 of the main text (McKinley et al., 2025).

#### S3.1 Full derivations of bounds

Section 3.1.1 in Part 1 of the main text (Williamson et al., 2025) describes a generic mechanism for deriving appropriate bounds on the embedded model discrepancy process for compartmental models. For illustration, here we follow that process through explicitly for all compartments in our model.

Starting with the absorbing states, consider first $Y_{t,D_H}$. We have that $Y_{t,D_H} = Y_{t-1,D_H} + Y'_{t,D_H}$, and from the model structure we can derive bounds

$$0 \leq Y'_{t,D_H} \leq Y_{t-1,H}.$$

Since the adjusted incidence $Y'_{t,D_H} = X'_{t,D_H} + \Delta'_{t,D_H}$, we can derive constraints on $\Delta'_{t,D_H}$ such that

$$-X'_{t,D_H} \leq \Delta'_{t,D_H} \leq Y_{t-1,H} - X'_{t,D_H}.$$

The adjusted count is therefore given by

$$Y_{t,D_H} = X_{t,D_H} + \Delta'_{t,D_H},$$

since

$$\begin{aligned} Y_{t,D_H} &= Y_{t-1,D_H} + Y'_{t,D_H}, \\ &= Y_{t-1,D_H} + X'_{t,D_H} + \Delta'_{t,D_H}, \\ &= X_{t,D_H} + \Delta'_{t,D_H} \qquad \text{since } X_{t,D_H} = Y_{t-1,D_H} + X'_{t,D_H}. \end{aligned}$$

This leads to the the form given in (S.1).

Now consider $Y_{t,D_I}$. From the model structure we can derive bounds for the MD as:

$$\begin{aligned} 0 \leq Y'_{t,D_I} &\leq Y_{t-1,I_1}, \\ 0 \leq X'_{t,D_I} + \Delta'_{t,D_I} &\leq Y_{t-1,I_1}, \\ -X'_{t,D_I} \leq \Delta'_{t,D_I} &\leq Y_{t-1,I_1} - X'_{t,D_I}. \end{aligned}$$

The adjusted counts are derived similarly to $Y_{t,D_H}$, leading to the the form given in (S.2).

Now consider $Y_{t,R_H}$. From the model structure we know that

$$\begin{aligned} Y'_{t,D_H} + Y'_{t,R_H} &\leq Y_{t-1,H}, \\ \Rightarrow 0 \leq Y'_{t,R_H} &\leq Y_{t-1,H} - Y'_{t,D_H}, \end{aligned}$$

and since $Y'_{t,R_H} = X'_{t,R_H} + \Delta'_{t,R_H}$ we can derive a constraint for $\Delta'_{t,R_H}$ conditional on the adjusted $Y'_{t,D_H}$, such that

$$-X'_{t,R_H} \leq \Delta'_{t,R_H} \leq Y_{t-1,H} - Y'_{t,D_H} - X'_{t,R_H}.$$

The adjusted counts are derived similarly to above, leading to the the form given in (S.3).

Now consider $Y_{t,R_I}$. From the model structure we can derive bounds for the MD as:

$$\begin{aligned} 0 \leq Y'_{t,R_I} &\leq Y_{t-1,I_2}, \\ 0 \leq X'_{t,R_I} + \Delta'_{t,R_I} &\leq Y_{t-1,I_2}, \\ -X'_{t,R_I} \leq \Delta'_{t,R_I} &\leq Y_{t-1,I_2} - X'_{t,R_I}. \end{aligned}$$

The adjusted counts are derived similarly to above, leading to the the form given in (S.4).

Now consider $Y_{t,R_A}$. From the model structure we can derive bounds for the MD as:

$$\begin{aligned} 0 \leq Y'_{t,R_A} &\leq Y_{t-1,A}, \\ 0 \leq X'_{t,R_A} + \Delta'_{t,R_A} &\leq Y_{t-1,A}, \\ -X'_{t,R_A} \leq \Delta'_{t,R_A} &\leq Y_{t-1,A} - X'_{t,R_A}. \end{aligned}$$

The adjusted counts are derived similarly to above, leading to the the form given in (S.5).

Now consider the non-absorbing states, starting with $Y_{t,H}$. We know from the model structure that

$$Y_{t,H} = Y_{t-1,H} + Y'_{t,H} - Y'_{t,D_H} - Y'_{t,R_H},$$

and we now place the model discrepancy on the state and *not* the incidence. Therefore,

$$Y_{t,H} = X_{t,H} + \Delta_{t,H}.$$

Hence

$$\begin{aligned}
Y_{t,H} &= Y_{t-1,H} + Y'_{t,H} - Y'_{t,D_H} - Y'_{t,R_H} \\
\Rightarrow Y'_{t,H} &= Y_{t,H} - Y_{t-1,H} + Y'_{t,D_H} + Y'_{t,R_H} \\
\Rightarrow Y'_{t,H} &= X_{t,H} + \Delta_{t,H} - Y_{t-1,H} + Y'_{t,D_H} + Y'_{t,R_H}.
\end{aligned}$$

Conditional on the adjusted counts simulated so far, we have that $0 \leq Y'_{t,H} \leq Y_{t-1,I_1} - Y'_{t,D_I}$, and as such we can derive bounds:

$$\begin{aligned}
0 \leq X_{t,H} + \Delta_{t,H} - Y_{t-1,H} + Y'_{t,D_H} + Y'_{t,R_H} &\leq Y_{t-1,I_1} - Y'_{t,D_I} \\
\Rightarrow -X_{t,H} + Y_{t-1,H} - Y'_{t,D_H} - Y'_{t,R_H} \leq \Delta_{t,H} &\leq Y_{t-1,I_1} - Y'_{t,D_I} - X_{t,H} + Y_{t-1,H} - Y'_{t,D_H} - Y'_{t,R_H},
\end{aligned}$$

leading to the the form given in (S.6).

Now consider $Y_{t,I_2}$. From the model structure we have that

$$Y_{t,I_2} = Y_{t-1,I_2} + Y'_{t,I_2} - Y'_{t,R_I},$$

and we place MD on the *counts*, such that:

$$Y_{t,I_2} = X_{t,I_2} + \Delta_{t,I_2}.$$

Hence:

$$\begin{aligned}
Y_{t,I_2} &= Y_{t-1,I_2} + Y'_{t,I_2} - Y'_{t,R_I} \\
\Rightarrow Y'_{t,I_2} &= Y_{t,I_2} - Y_{t-1,I_2} + Y'_{t,R_I} \\
\Rightarrow Y'_{t,I_2} &= X_{t,I_2} + \Delta_{t,I_2} - Y_{t-1,I_2} + Y'_{t,R_I}
\end{aligned}$$

Conditional on the adjusted counts simulated so far, we have that $0 \leq Y'_{t,I_2} \leq Y_{t-1,I_1} - Y'_{t,D_I} - Y'_{t,H}$, and as such we can derive bounds:

$$\begin{aligned}
0 \leq Y'_{t,I_2} &\leq Y_{t-1,I_1} - Y'_{t,D_I} - Y'_{t,H} \\
\Rightarrow 0 \leq X_{t,I_2} + \Delta_{t,I_2} - Y_{t-1,I_2} + Y'_{t,R_I} &\leq Y_{t-1,I_1} - Y'_{t,D_I} - Y'_{t,H} \\
\Rightarrow -X_{t,I_2} + Y_{t-1,I_2} - Y'_{t,R_I} \leq \Delta_{t,I_2} &\leq Y_{t-1,I_1} - Y'_{t,D_I} - Y'_{t,H} - X_{t,I_2} + Y_{t-1,I_2} - Y'_{t,R_I},
\end{aligned}$$

leading to the the form given in (S.7).

Now consider $Y_{t,I_1}$. From the model structure we have that

$$Y_{t,I_1} = Y_{t-1,I_1} + Y'_{t,I_1} - Y'_{t,I_2} - Y'_{t,D_I} - Y'_{t,H},$$

and we place MD on the *counts*, such that:

$$Y_{t,I_1} = X_{t,I_1} + \Delta_{t,I_1}$$

Hence:

$$\begin{aligned}
Y_{t,I_1} &= Y_{t-1,I_1} + Y'_{t,I_1} - Y'_{t,I_2} - Y'_{t,D_I} - Y'_{t,H} \\
Y'_{t,I_1} &= Y_{t,I_1} - Y_{t-1,I_1} + Y'_{t,I_2} + Y'_{t,D_I} + Y'_{t,H}
\end{aligned}$$

and since $0 \leq Y'_{t,I_1} \leq Y_{t-1,P}$ we can derive bounds:

$$\begin{aligned}
0 \leq Y'_{t,I_1} &\leq Y_{t-1,P}, \\
0 \leq Y_{t,I_1} - Y_{t-1,I_1} + Y'_{t,I_2} + Y'_{t,D_I} + Y'_{t,H} &\leq Y_{t-1,P}, \\
0 \leq X_{t,I_1} + \Delta_{t,I_1} - Y_{t-1,I_1} + Y'_{t,I_2} + Y'_{t,D_I} + Y'_{t,H} &\leq Y_{t-1,P} \\
-X_{t,I_1} + Y_{t-1,I_1} - Y'_{t,I_2} - Y'_{t,D_I} - Y'_{t,H} \leq \Delta_{t,I_1} &\leq Y_{t-1,P} - X_{t,I_1} + Y_{t-1,I_1} - Y'_{t,I_2} - Y'_{t,D_I} - Y'_{t,H},
\end{aligned}$$

leading to the the form given in (S.8).

Now consider $Y_{t,P}$. From the model structure we have that

$$Y_{t,P} = Y_{t-1,P} + Y'_{t,P} - Y'_{t,I_1}$$

with MD placed on the *counts* as

$$Y_{t,P} = X_{t,P} + \Delta_{t,P}.$$

Since $0 \leq Y'_{t,P} \leq Y_{t-1,E}$, we can derive bounds

$$\begin{aligned}
0 &\leq Y'_{t,P} \leq Y_{t-1,E}, \\
0 &\leq Y_{t,P} - Y_{t-1,P} + Y'_{t,I_1} \leq Y_{t-1,E}, \\
0 &\leq X_{t,P} + \Delta_{t,P} - Y_{t-1,P} + Y'_{t,I_1} \leq Y_{t-1,E}, \\
-X_{t,P} + Y_{t-1,P} - Y'_{t,I_1} &\leq \Delta_{t,P} \leq Y_{t-1,E} - X_{t,P} + Y_{t-1,P} - Y'_{t,I_1},
\end{aligned}$$

leading to the the form given in (S.9).

Now consider $Y_{t,A}$. From the model structure we have that

$$Y_{t,A} = Y_{t-1,A} + Y'_{t,A} - Y'_{t,R_A}$$

and place MD on the *counts*, such that

$$Y_{t,A} = X_{t,A} + \Delta_{t,A}$$

Conditional on the adjusted counts simulated so far, we have that $0 \leq Y'_{t,A} \leq Y_{t-1,E} - Y'_{t,P}$, and thus we can derive bounds:

$$\begin{aligned}
0 &\leq Y'_{t,A} \leq Y_{t-1,E} - Y'_{t,P} \\
0 &\leq Y_{t,A} - Y_{t-1,A} + Y'_{t,R_A} \leq Y_{t-1,E} - Y'_{t,P} \\
0 &\leq X_{t,A} + \Delta_{t,A} - Y_{t-1,A} + Y'_{t,R_A} \leq Y_{t-1,E} - Y'_{t,P} \\
-X_{t,A} + Y_{t-1,A} - Y'_{t,R_A} &\leq \Delta_{t,A} \leq Y_{t-1,E} - Y'_{t,P} - X_{t,A} + Y_{t-1,A} - Y'_{t,R_A}
\end{aligned}$$

leading to the the form given in (S.10).

Now consider $Y_{t,E}$. From the model structure we have that

$$Y_{t,E} = Y_{t-1,E} + Y'_{t,E} - Y'_{t,P} - Y'_{t,A}$$

and we place MD on the *count*, such that

$$Y_{t,E} = X_{t,E} + \Delta_{t,E}$$

Since $0 \leq Y'_{t,E} \leq Y_{t-1,S}$ we can derive bounds:

$$\begin{aligned}
0 &\leq Y'_{t,E} \leq Y_{t-1,S}, \\
0 &\leq Y_{t,E} - Y_{t-1,E} + Y'_{t,P} + Y'_{t,A} \leq Y_{t-1,S}, \\
0 &\leq X_{t,E} + \Delta_{t,E} - Y_{t-1,E} + Y'_{t,P} + Y'_{t,A} \leq Y_{t-1,S} \\
-X_{t,E} + Y_{t-1,E} - Y'_{t,P} - Y'_{t,A} &\leq \Delta_{t,E} \leq Y_{t-1,S} - X_{t,E} + Y_{t-1,E} - Y'_{t,P} - Y'_{t,A},
\end{aligned}$$

leading to the the form given in (S.11). Finally we have

$$Y_{t,S} = Y_{t-1,S} - Y'_{t,E}.$$

### S4 Sampling from conditional distributions

#### S4.1 Sampling from conditional simulator

It turns out that we can sample from $\pi\left(\boldsymbol{X}_t^{-z} \mid \boldsymbol{X}_t^z, \boldsymbol{Y}_{t-1}\right)$ *directly from the simulator*, by adjusting some parameters and inputs slightly. In the subsequent discussion we present the case for sampling from a single region and age-class, and for clarity we drop the $s$ and $a$ subscripts.

If we first consider

$$\left(X'_{t,D_H}, X'_{t,R_H}, \cdot \mid \boldsymbol{Y}_{t-1}\right) \sim \text{Multinomial}\left(Y_{t-1,H}, \boldsymbol{p}\right), \tag{S.12}$$

where $\boldsymbol{p} = \left(p_H p_{HD}, p_H\left(1-p_{HD}\right), 1-p_H\right)$. (Here the $\cdot$ represents those individuals who do not transition between states.) From this we have that

$$\left(X'_{t,R_H} \mid X'_{t,D_H}, \boldsymbol{Y}_{t-1}\right) \sim \text{Binomial}\left(Y_{t-1,H} - X'_{t,D_H}, \frac{p_H\left(1-p_{HD}\right)}{1-p_H p_{HD}}\right). \tag{S.13}$$

Sampling from (S.13) is equivalent to sampling from

$$\left(D, X'_{t,R_H}, \cdot \mid X'_{t,D_H}, \boldsymbol{Y}_{t-1}\right) \sim \text{Multinomial}\left(Y_{t-1,H} - X'_{t,D_H}, \boldsymbol{p}^*\right), \tag{S.14}$$

where $\boldsymbol{p}^* = \left(0, \frac{p_H(1-p_{HD})}{1-p_H p_{HD}}, \frac{1-p_H}{1-p_H p_{HD}}\right)$ and $D=0$ is a dummy variable. Similarly, if we consider

$$\left(X'_{t,H}, X'_{t,I_2}, X'_{t,D_I}, \cdot \mid \boldsymbol{Y}_{t-1}\right) \sim \text{Multinomial}\left(Y_{t-1,I_1}, \boldsymbol{q}\right), \tag{S.15}$$

where $\boldsymbol{q} = \left(p_{I_1} p_{I_1 H}, p_{I_1}\left(1 - p_{I_1 H} - p_{I_1 D}\right), p_{I_1} p_{I_1 D}, 1 - p_{I_1}\right)$, then we can derive the conditional:

$$\left(X'_{t,H}, X'_{t,I_2}, \cdot \mid X'_{t,D_I}, \boldsymbol{Y}_{t-1}\right) \sim \text{Multinomial}\left(Y_{t-1,I_1} - X'_{t,D_I}, \boldsymbol{q}'\right), \tag{S.16}$$

where $\boldsymbol{q}' = \left(\frac{p_{I_1} p_{I_1 H}}{1-p_{I_1} p_{I_1 D}}, \frac{p_{I_1}\left(1-p_{I_1 H}-p_{I_1 D}\right)}{1-p_{I_1} p_{I_1 D}}, \frac{1-p_{I_1}}{1-p_{I_1} p_{I_1 D}}\right)$. Sampling from (S.16) is equivalent to sampling from

$$\left(X'_{t,H}, X'_{t,I_2}, D, \cdot \mid X'_{t,D_I}, \boldsymbol{Y}_{t-1}\right) \sim \text{Multinomial}\left(Y_{t-1,I_1} - X'_{t,D_I}, \boldsymbol{q}^*\right), \tag{S.17}$$

where $\boldsymbol{q}^* = \left(\frac{p_{I_1} p_{I_1 H}}{1-p_{I_1} p_{I_1 D}}, \frac{p_{I_1}\left(1-p_{I_1 H}-p_{I_1 D}\right)}{1-p_{I_1} p_{I_1 D}}, 0, \frac{1-p_{I_1}}{1-p_{I_1} p_{I_1 D}}\right)$ and $D=0$ is a dummy variable. Hence we can use the existing simulator code to sample from the conditional simulator by just adjusting the inputs and parameters.

#### S4.2 Sampling from conditional model discrepancy distribution

The conditional model discrepancy distribution is:

$$\begin{aligned} f\left(\boldsymbol{Y}_t^{-z} \mid \boldsymbol{Y}_t^z, \boldsymbol{X}_t^{-z}, \boldsymbol{Y}_{t-1}\right) = {} & p\left(\boldsymbol{Y}'_{t,R_H} \mid \boldsymbol{Y}'_{t,D_H}, \boldsymbol{X}_t, \boldsymbol{Y}_{t-1}\right) p\left(\boldsymbol{Y}'_{t,R_I} \mid \boldsymbol{X}_t, \boldsymbol{Y}_{t-1}\right) p\left(\boldsymbol{Y}'_{t,R_A} \mid \boldsymbol{X}_t, \boldsymbol{Y}_{t-1}\right) \\ & \times p\left(\boldsymbol{Y}_{t,H} \mid \boldsymbol{Y}'_{t,D_H}, \boldsymbol{Y}'_{t,R_H}, \boldsymbol{Y}'_{t,D_I}, \boldsymbol{X}_t, \boldsymbol{Y}_{t-1}\right) \\ & \times p\left(\boldsymbol{Y}_{t,I_2} \mid \boldsymbol{Y}'_{t,H}, \boldsymbol{Y}'_{t,D_I}, \boldsymbol{Y}'_{t,R_I}, \boldsymbol{X}_t, \boldsymbol{Y}_{t-1}\right) \\ & \times p\left(\boldsymbol{Y}_{t,I_1} \mid \boldsymbol{Y}'_{t,I_2}, \boldsymbol{Y}'_{t,D_I}, \boldsymbol{Y}'_{t,H}, \boldsymbol{X}_t, \boldsymbol{Y}_{t-1}\right) p\left(\boldsymbol{Y}_{t,P} \mid \boldsymbol{Y}'_{t,I_1}, \boldsymbol{X}_t, \boldsymbol{Y}_{t-1}\right) \\ & \times p\left(\boldsymbol{Y}_{t,A} \mid \boldsymbol{Y}'_{t,R_A}, \boldsymbol{X}_t, \boldsymbol{Y}_{t-1}\right) p\left(\boldsymbol{Y}_{t,E} \mid \boldsymbol{Y}'_{t,P}, \boldsymbol{Y}'_{t,A}, \boldsymbol{X}_t, \boldsymbol{Y}_{t-1}\right), \end{aligned} \tag{S.18}$$

and here all the component distributions in (S.18) are independent over space and age given $\boldsymbol{Y}_t^z$, $\boldsymbol{X}_t^{-z}$ and $\boldsymbol{Y}_{t-1}$ and thus are straightforward to sample from.

#### S5 Simulation study parameters

| Parameter | Value |
|---|---|
| $R_0$ | 4 |
| $\nu_A$ | 0.37 |
| $T_E$ | 0.91 |
| $T_P$ | 1.6 |
| $T_{I_1}$ | 3.8 |
| $T_{I_2}$ | 0.24 |
| $\alpha_{EP}$ | -4.3 |
| $\alpha_{I_1H}$ | -3.6 |
| $\alpha_{I_1D}$ | -8.1 |
| $\alpha_{HD}$ | -4.4 |
| $\eta$ | 0.038 |
| $\eta_I$ | 0.74 |
| $\eta_H$ | 1.4 |
| $\alpha_{TH}$ | 0.47 |
| $\eta_{TH}$ | 0.027 |
| $\beta_s$ | 0.27 |
| $p_{\text{move}}$ | 0.49 |
| $\alpha_{MD}$ | 0.24 |
| $t_{\text{MD, East of England}}$ | 20 |
| $t_{\text{MD, East Midlands}}$ | 36 |
| $t_{\text{MD, London}}$ | 26 |
| $t_{\text{MD, North-East}}$ | 25 |
| $t_{\text{MD, North-West}}$ | 18 |
| $t_{\text{MD, South-East}}$ | 34 |
| $t_{\text{MD, South-West}}$ | 29 |
| $t_{\text{MD, West Midlands}}$ | 30 |
| $t_{\text{MD, Yorkshire and the Humber}}$ | 5 |

Table S1: Parameters chosen for simulation study (to 2 significant figures)

### S6 Additional figures

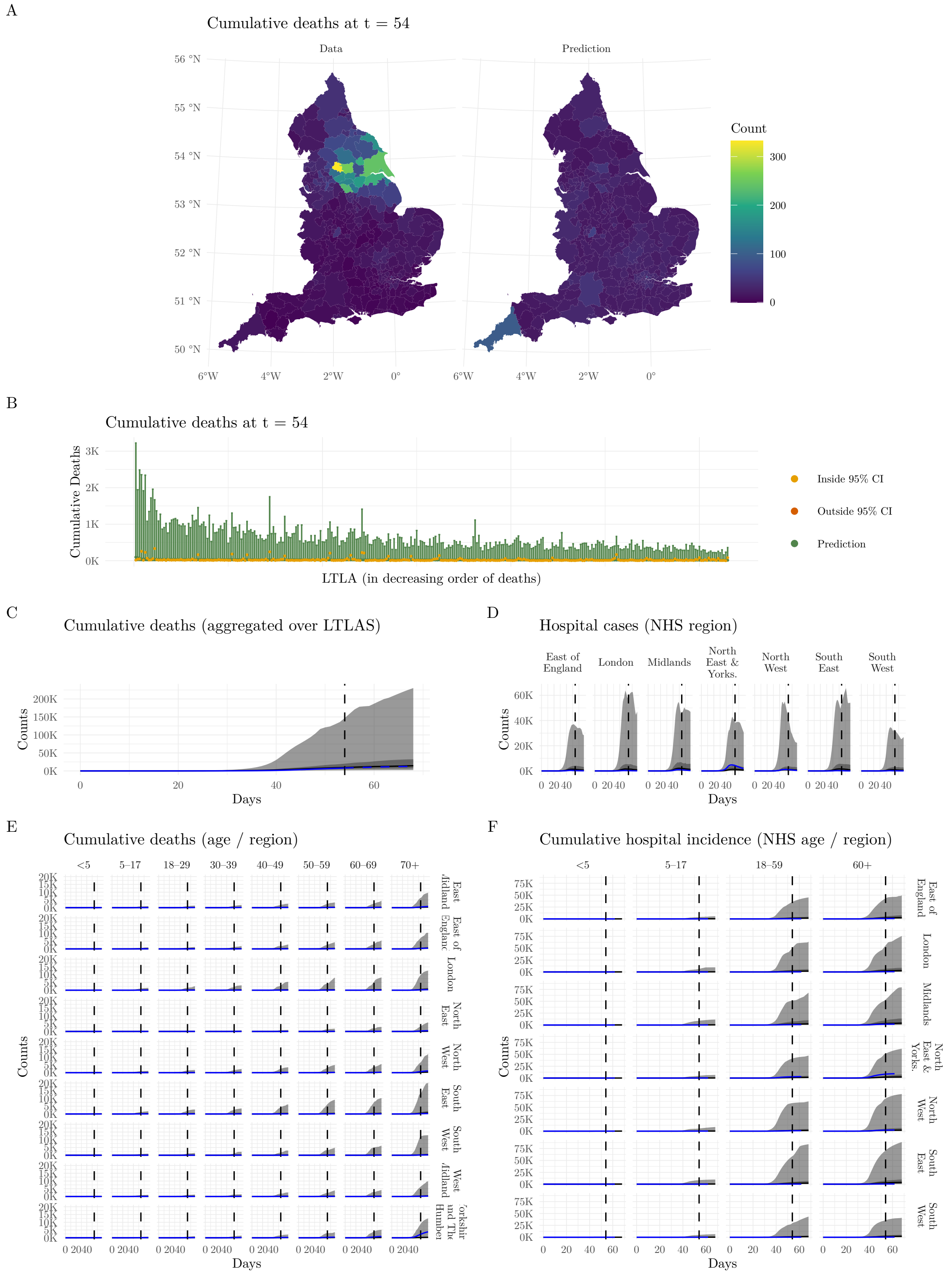


Figure S1: Particle trajectory plots across the ensemble of design points at Wave 1 for the simulated outbreak. A) Spatial (LTLA-level) plots of the mean number of deaths by day 54, for the data and the fitted model. B) Cumulative deaths at day 54 within each LTLA (ranked in decreasing order of predicted deaths). C) Cumulative deaths over time, aggregated over the 338 LTLAs. D) Hospital cases over time in each NHS region. E) Cumulative deaths over time in each age/region category. F) Cumulative hospital incidence over time by each NHS age/region category. In plot B) the green points are the predicted ensemble means, and the error bars are the 95% prediction intervals. The yellow and red points are the observed data coloured by whether they lie inside and outside of the prediction intervals respectively. In plots C–F, the blue dashed lines correspond to the observed data and the black solid lines to the mean trajectories from the particles taken across the ensemble. The ribbons correspond to 50% and 95% prediction intervals. A vertical dashed line corresponds to the end point of the observed data, such that trajectories before the line are generated from the particle filter, and trajectories to the right of the line are simulated forecasts from the model.

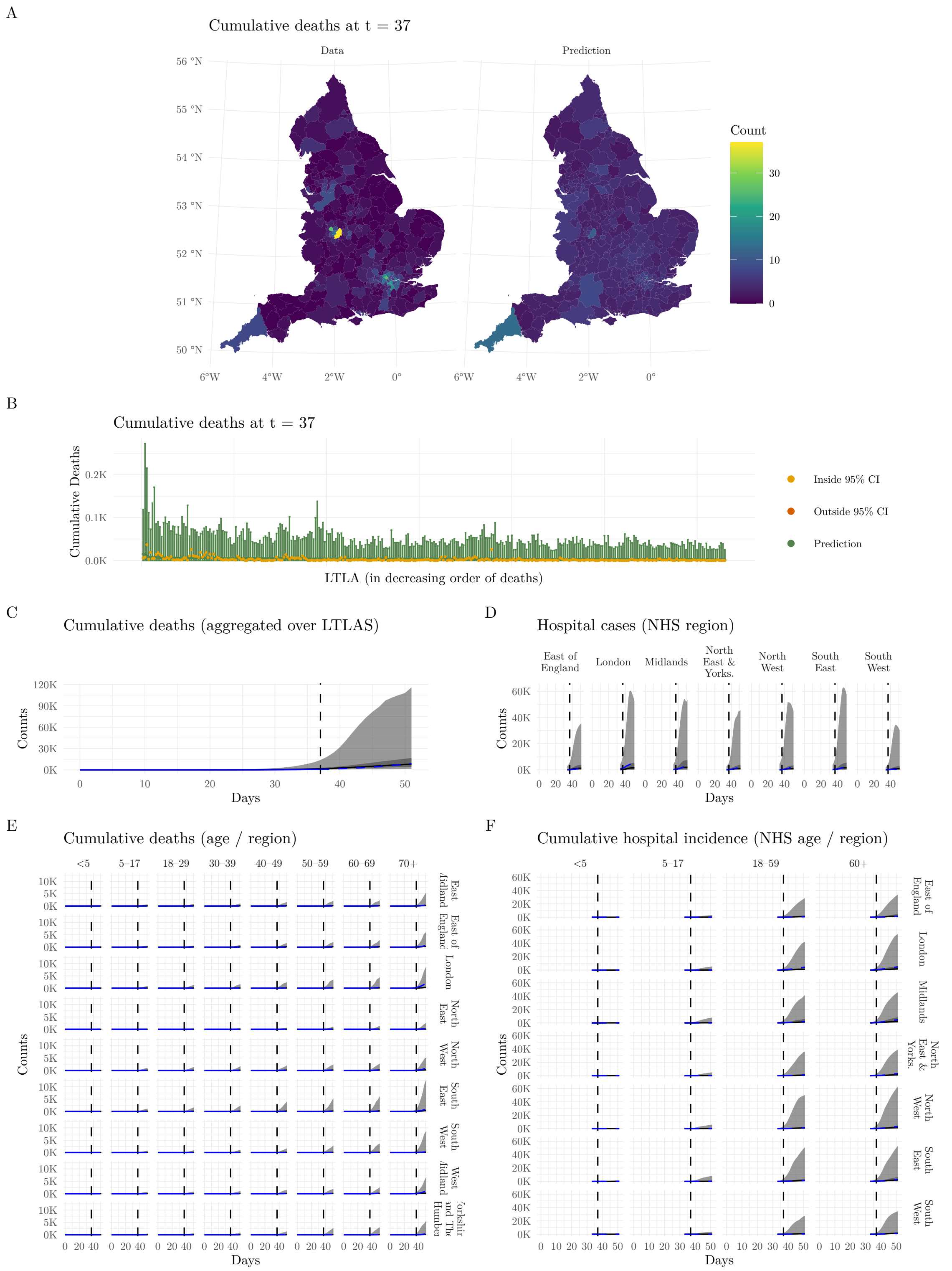


Figure S2: Particle trajectory plots across the ensemble of design points at Wave 1 for the real UK data up to the first lockdown. A) Spatial (LTLA-level) plots of the mean number of deaths by day 37, for the data and the fitted model. B) Cumulative deaths at day 37 within each LTLA (ranked in decreasing order of predicted deaths). C) Cumulative deaths over time, aggregated over the 338 LTLAs. D) Hospital cases over time in each NHS region. E) Cumulative deaths over time in each age/region category. F) Cumulative hospital incidence over time by each NHS age/region category. In plot B) the green points are the predicted ensemble means, and the error bars are the 95% prediction intervals. The yellow and red points are the observed data coloured by whether they lie inside and outside of the prediction intervals respectively. In plots C–F, the blue dashed lines correspond to the observed data and the black solid lines to the mean trajectories from the particles taken across the ensemble. The ribbons correspond to 50% and 95% prediction intervals. A vertical dashed line corresponds to the end point of the observed data, such that trajectories before the line are generated from the particle filter, and trajectories to the right of the line are simulated forecasts from the model.

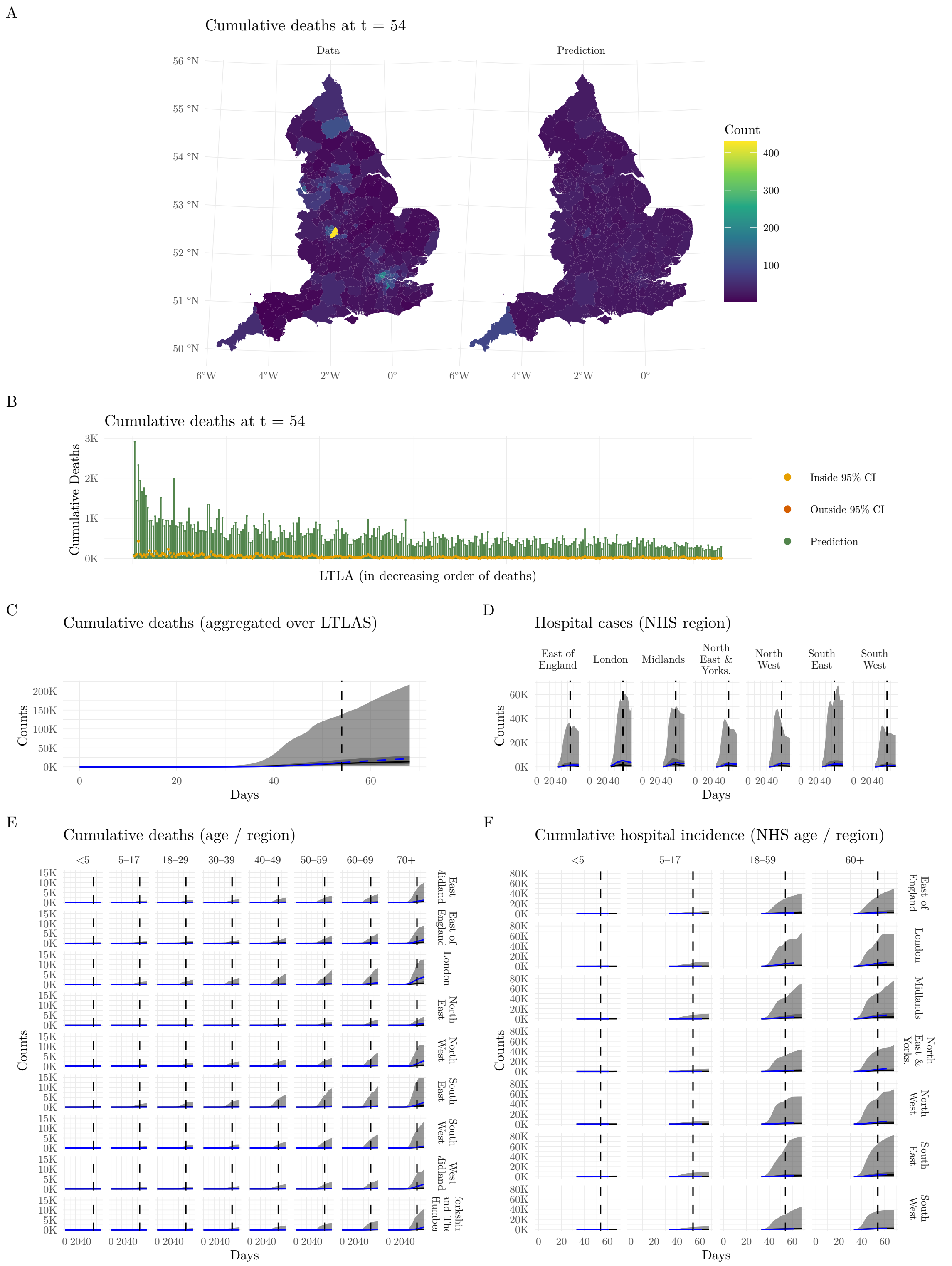


Figure S3: Particle trajectory plots across the ensemble of design points at Wave 1 for the real UK data beyond the first lockdown. A) Spatial (LTLA-level) plots of the mean number of deaths by day 54, for the data and the fitted model. B) Cumulative deaths at day 54 within each LTLA (ranked in decreasing order of predicted deaths). C) Cumulative deaths over time, aggregated over the 338 LTLAs. D) Hospital cases over time in each NHS region. E) Cumulative deaths over time in each age/region category. F) Cumulative hospital incidence over time by each NHS age/region category. In plot B) the green points are the predicted ensemble means, and the error bars are the 95% prediction intervals. The yellow and red points are the observed data coloured by whether they lie inside and outside of the prediction intervals respectively. In plots C–F, the blue dashed lines correspond to the observed data and the black solid lines to the mean trajectories from the particles taken across the ensemble. The ribbons correspond to 50% and 95% prediction intervals. A vertical dashed line corresponds to the end point of the observed data, such that trajectories before the line are generated from the particle filter, and trajectories to the right of the line are simulated forecasts from the model.

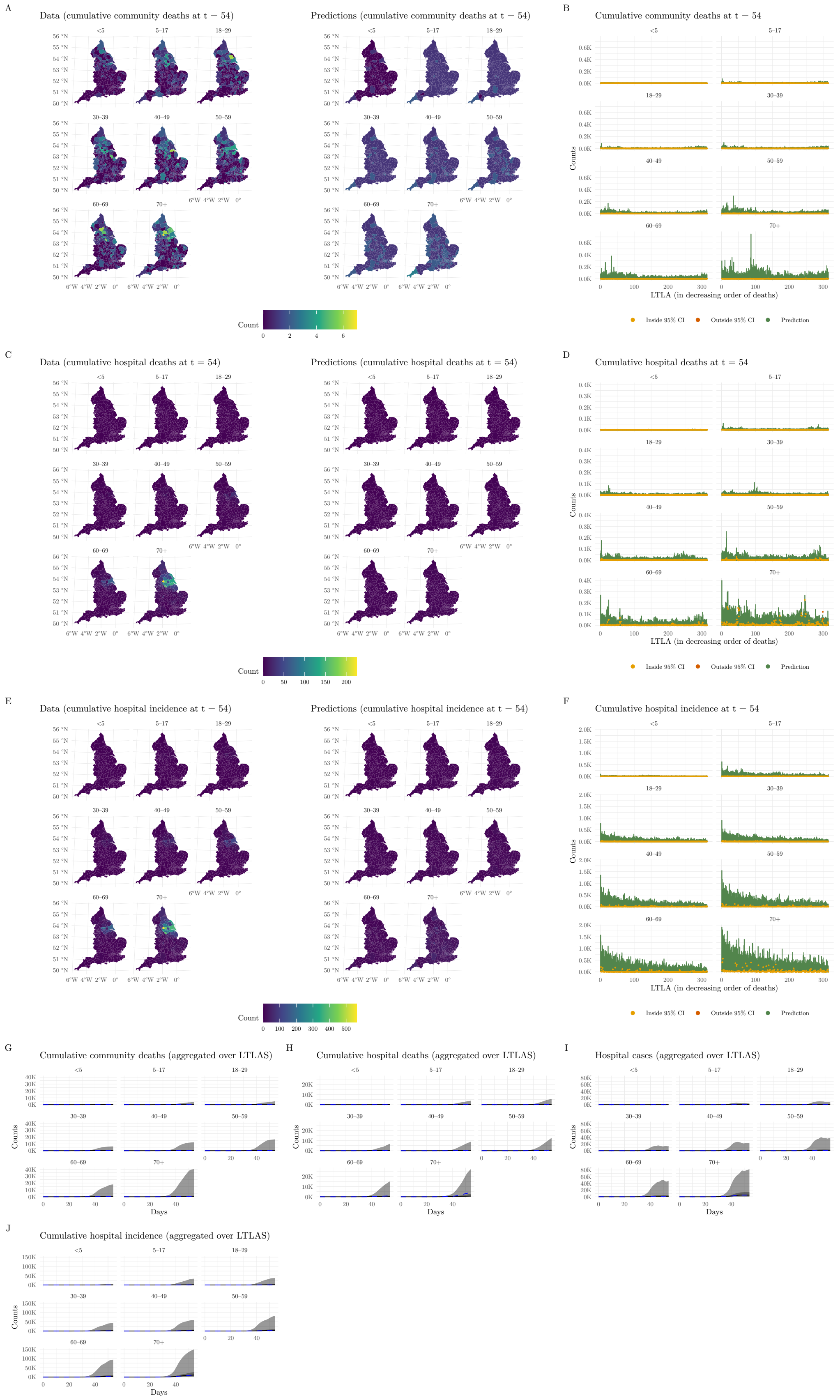


Figure S4: Particle trajectory plots across the ensemble of design points at Wave 1 for the simulated outbreak assuming data are available at the resolution of the model. Spatial (LTLA-level) plots of the data and the fitted model, are given for A) the mean number of community deaths, C) the mean number of hospital deaths, and C) the cumulative hospital incidence by day 54. Similarly, predicted and observed counts within each LTLA (ranked in decreasing order of predicted deaths) are given for C) the mean number of community deaths, D) the mean number of hospital deaths, and F) the cumulative hospital incidence by day 54 in each age category. Counts over time, by each age category aggregated over the 338 LTLAs are given for G) cumulative community deaths, H) cumulative hospital deaths, I) hospital cases and J) cumulative hospital incidence. In plots B), D) and F) the green points are the predicted ensemble means, and the error bars are the 95% prediction intervals. The yellow and red points are the observed data coloured by whether they lie inside and outside of the prediction intervals respectively. In plots G–J, the blue dashed lines correspond to the observed data and the black solid lines to the mean trajectories from the particles taken across the ensemble. The ribbons correspond to 50% and 95% prediction intervals.

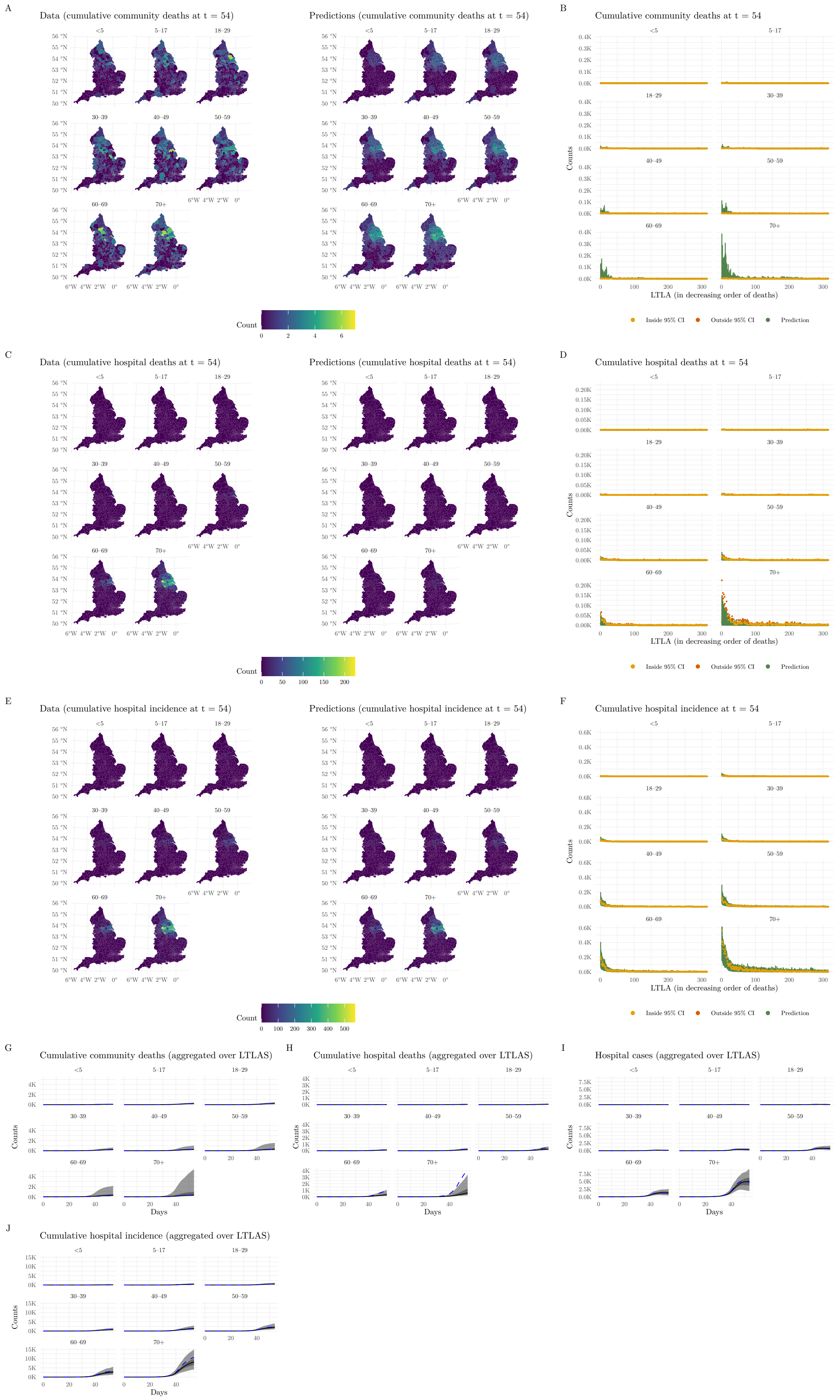


Figure S5: Particle trajectory plots across the ensemble of design points at Wave 10 for the simulated outbreak assuming data are available at the resolution of the model. Spatial (LTLA-level) plots of the data and the fitted model, are given for A) the mean number of community deaths, C) the mean number of hospital deaths, and C) the cumulative hospital incidence by day 54. Similarly, predicted and observed counts within each LTLA (ranked in decreasing order of predicted deaths) are given for C) the mean number of community deaths, D) the mean number of hospital deaths, and F) the cumulative hospital incidence by day 54 in each age category. Counts over time, by each age category aggregated over the 338 LTLAs are given for G) cumulative community deaths, H) cumulative hospital deaths, I) hospital cases and J) cumulative hospital incidence. In plots B), D) and F) the green points are the predicted ensemble means, and the error bars are the 95% prediction intervals. The yellow and red points are the observed data coloured by whether they lie inside and outside of the prediction intervals respectively. In plots G–J, the blue dashed lines correspond to the observed data and the black solid lines to the mean trajectories from the particles taken across the ensemble. The ribbons correspond to 50% and 95% prediction intervals.

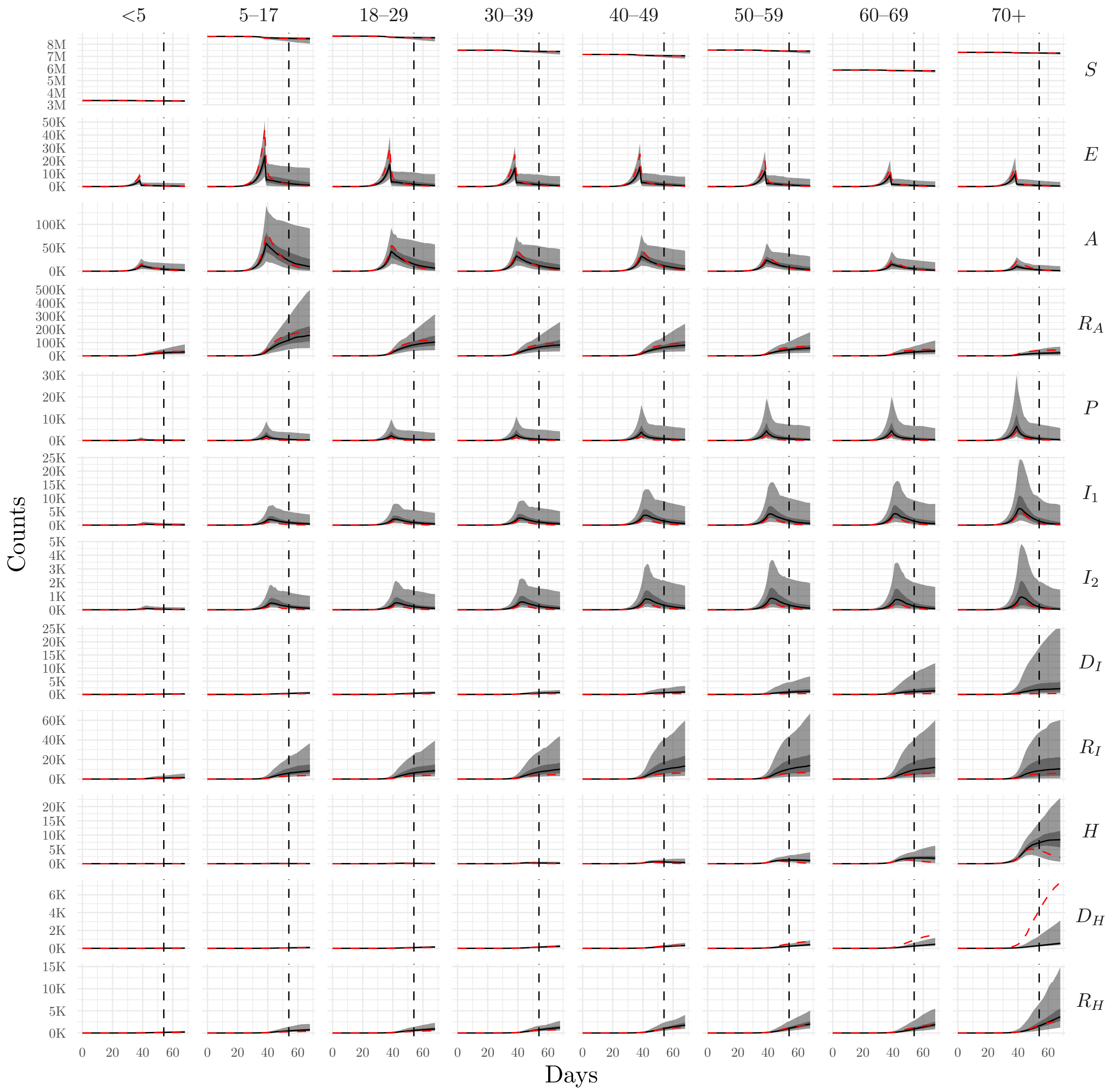


Figure S6: Particle trajectory plots for the hidden states across the ensemble of design points at Wave 10 for the simulated outbreak. The red lines correspond to the true trajectories, the black lines to the mean trajectories from the particles taken across the ensemble, and the ribbons correspond to 50% and 95% prediction intervals. A vertical dashed line corresponds to the end point of the observed data, such that trajectories before the line are generated from the particle filter, and trajectories to the right of the line are simulated forecasts from the model.

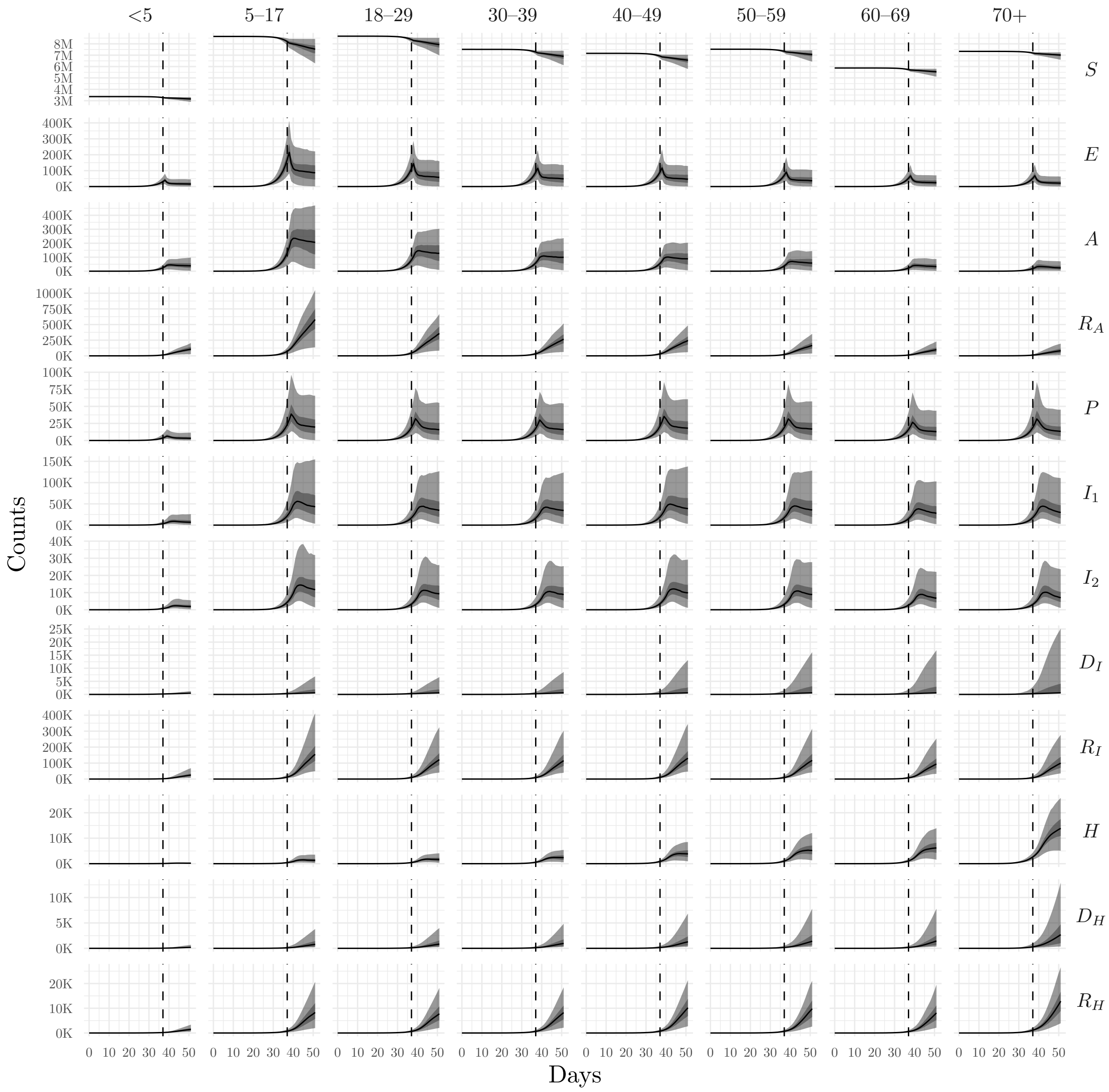


Figure S7: Particle trajectory plots for the hidden states across the ensemble of design points at Wave 10 for the real UK data up to the first lockdown. The black lines are the mean trajectories from the particles taken across the ensemble, and the ribbons correspond to 50% and 95% prediction intervals. A vertical dashed line corresponds to the end point of the observed data, such that trajectories before the line are generated from the particle filter, and trajectories to the right of the line are simulated forecasts from the model.

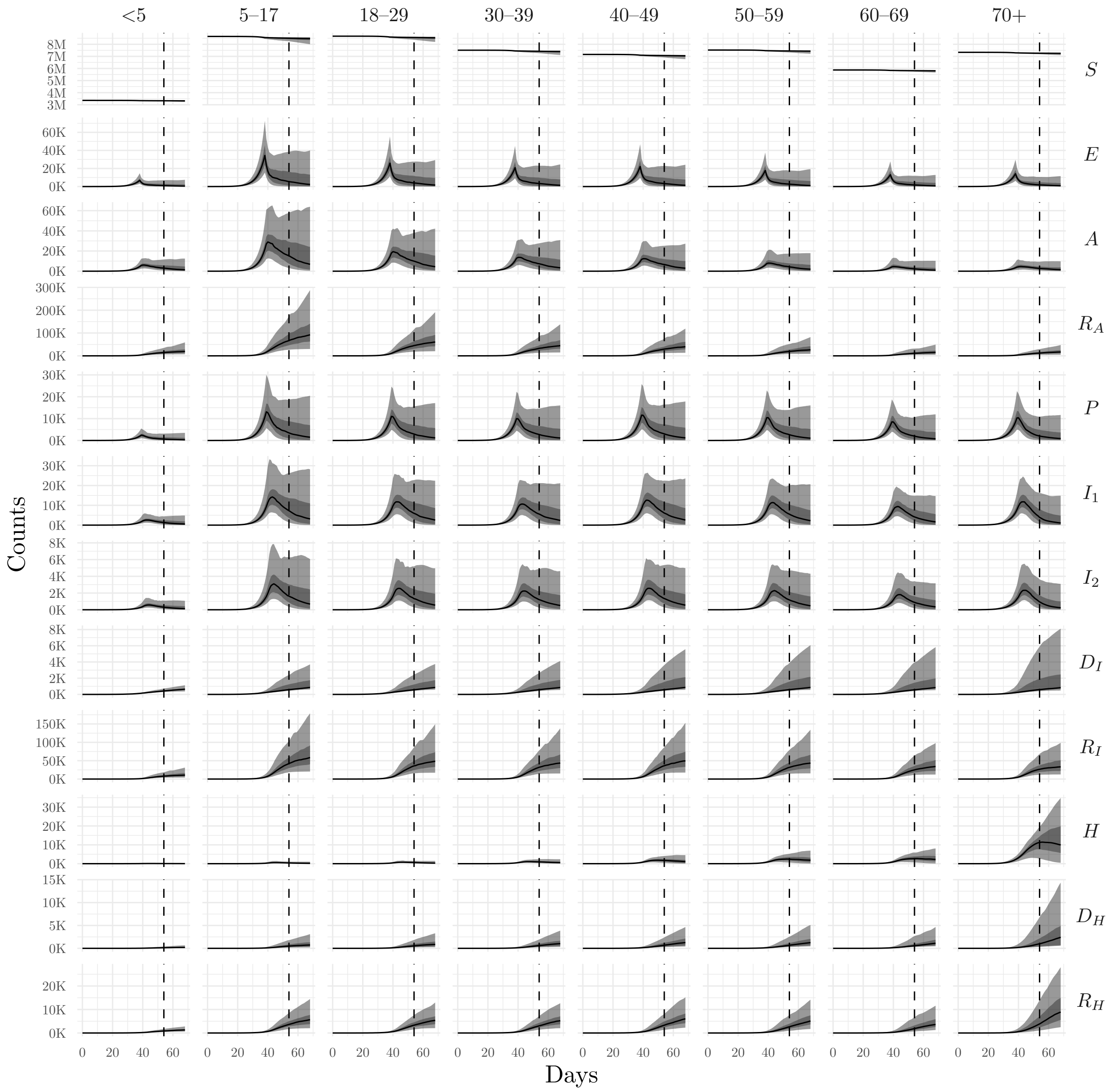


Figure S8: Particle trajectory plots for the hidden states across the ensemble of design points at Wave 10 for the real UK data beyond the first lockdown. The black lines are the mean trajectories from the particles taken across the ensemble, and the ribbons correspond to 50% and 95% prediction intervals. A vertical dashed line corresponds to the end point of the observed data, such that trajectories before the line are generated from the particle filter, and trajectories to the right of the line are simulated forecasts from the model.

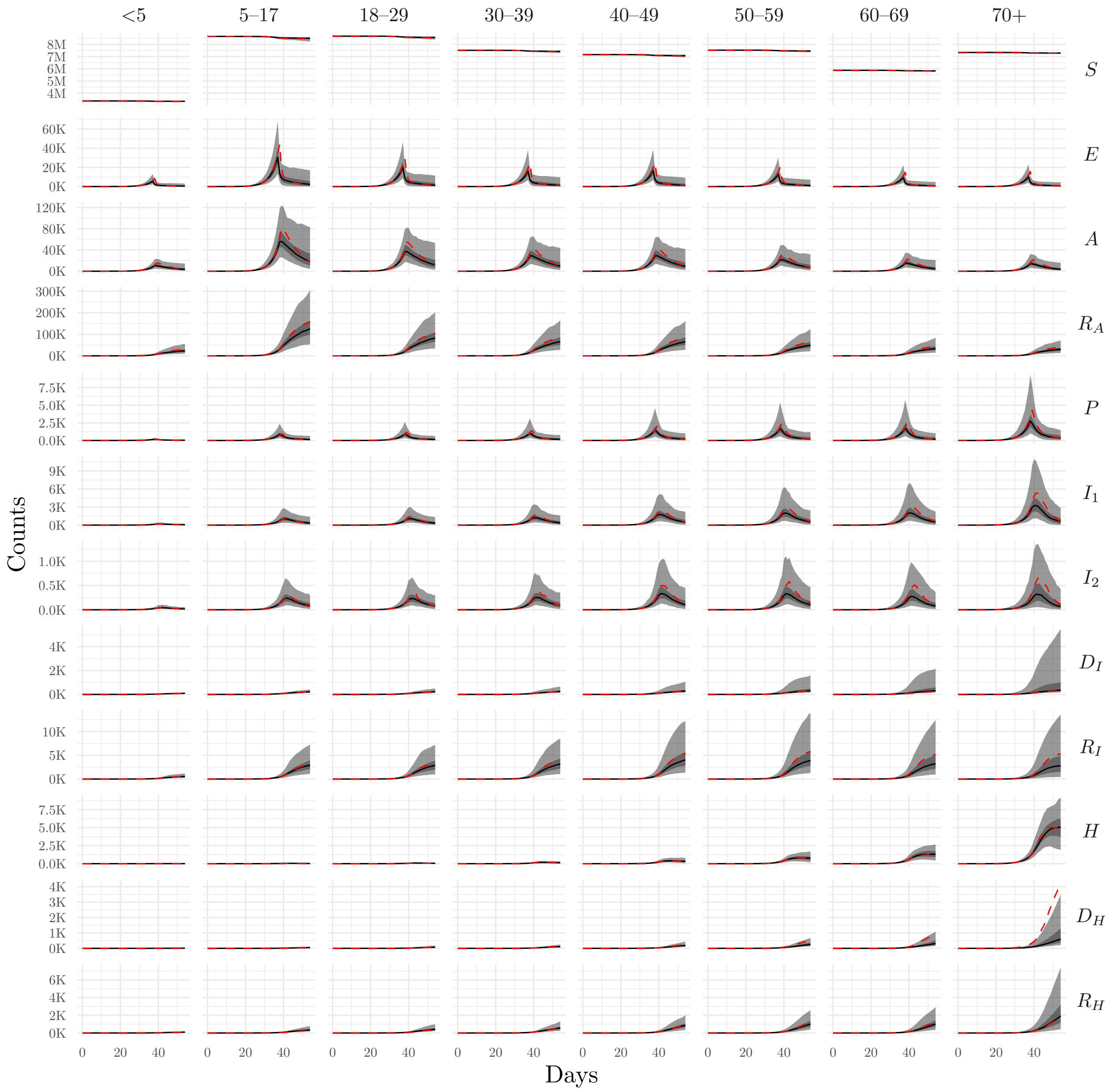


Figure S9: Particle trajectory plots for the hidden states across the ensemble of design points at Wave 10 for the simulated outbreak assuming data are available at the resolution of the model. The red lines correspond to the true trajectories, the black lines to the mean trajectories from the particles taken across the ensemble, and the ribbons correspond to 50% and 95% prediction intervals.

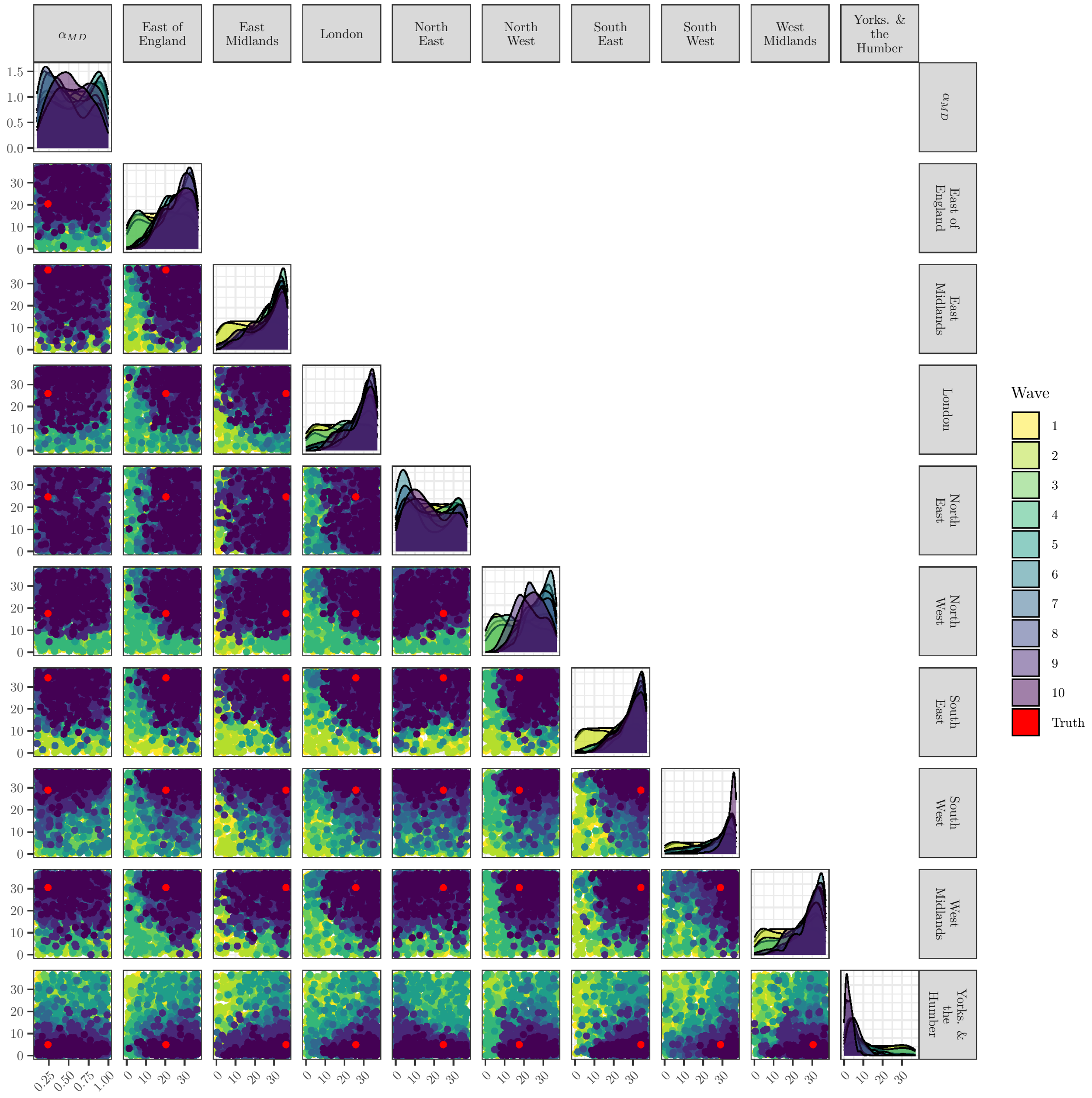


Figure S10: Inputs for Waves 1–10 for the $t_{r,\text{MD}}$ parameters from a model fitted to a simulated outbreak. The red points correspond to the true parameter values.

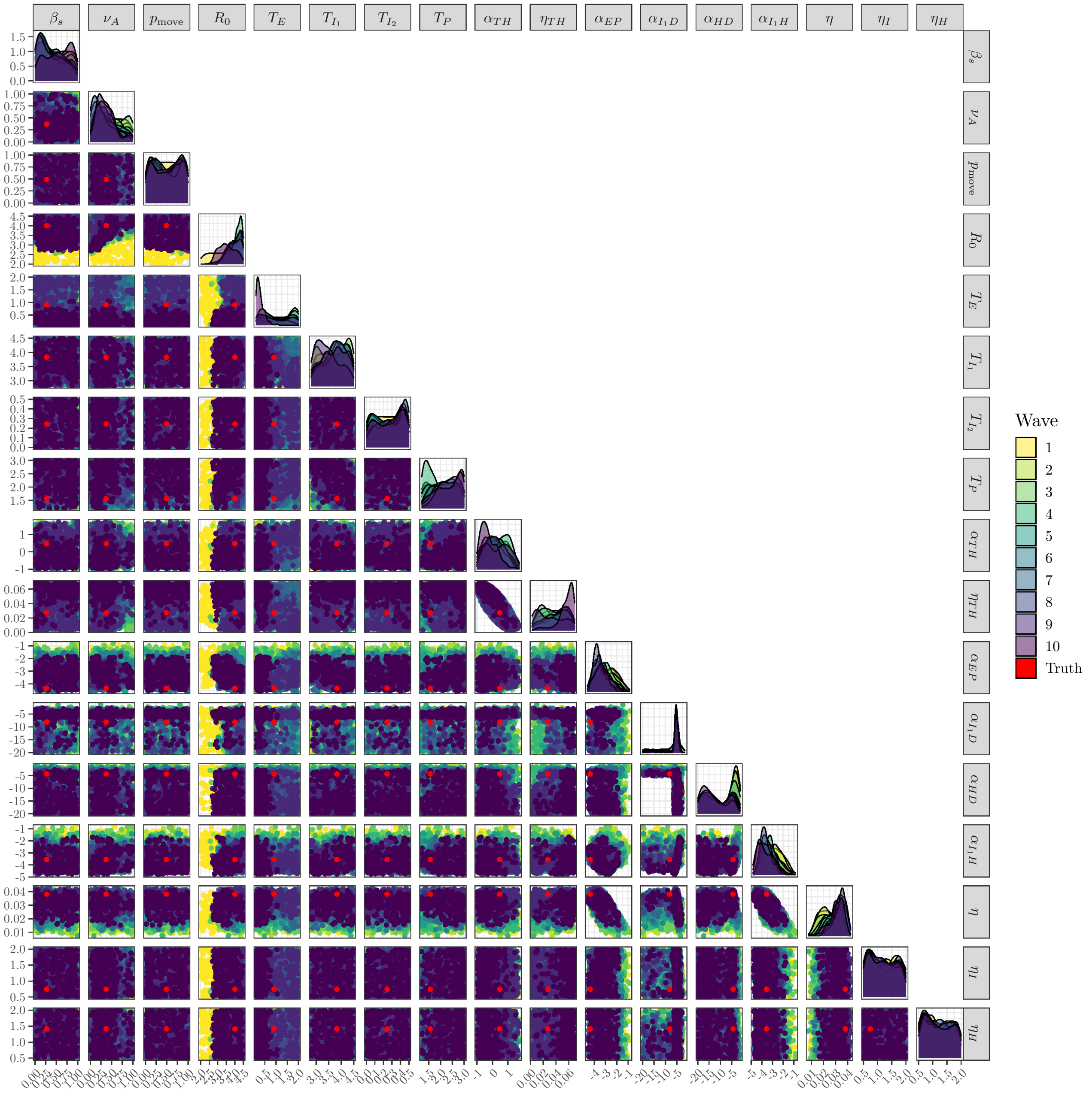


Figure S11: Inputs for Waves 1–10 for all parameters except the $t_{r,\mathrm{MD}}$ parameters from a model fitted to a simulated outbreak. The red points correspond to the true parameter values.

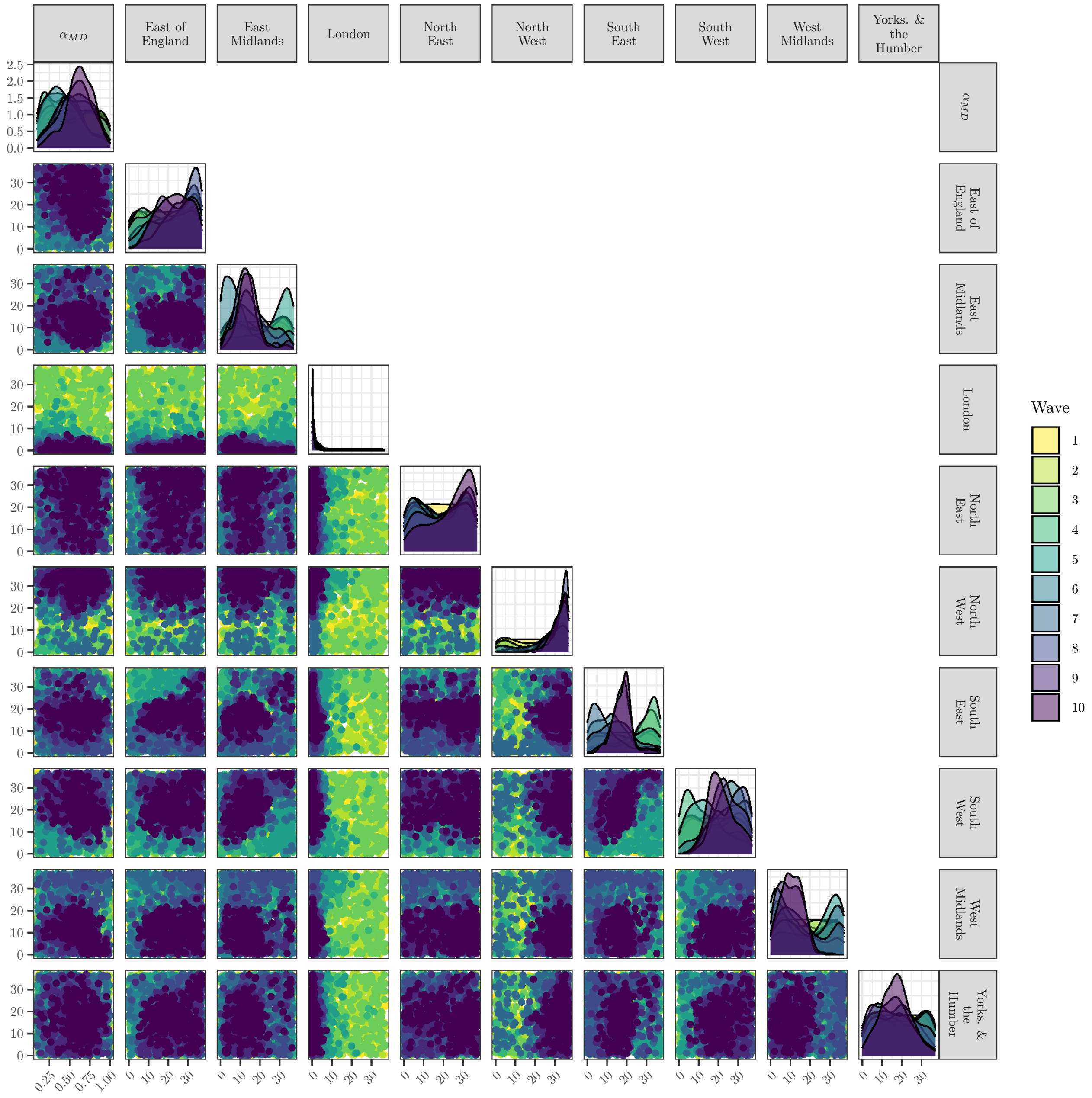


Figure S12: Inputs for Waves 1–10 for the $t_{r,\mathrm{MD}}$ parameters from a model fitted to the real UK data up to the first lockdown.

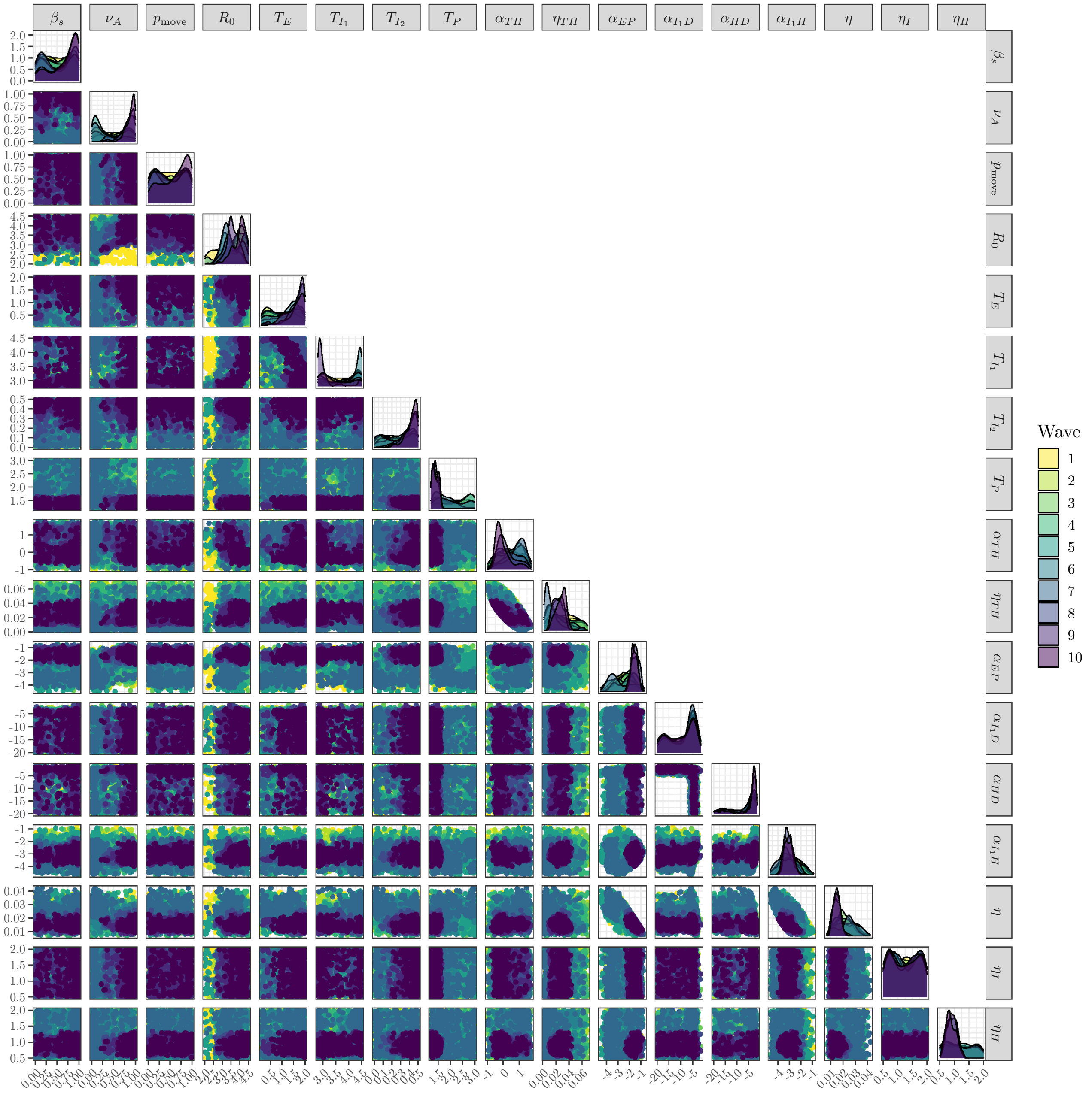


Figure S13: Inputs for Waves 1–10 for all parameters except the $t_{r,\text{MD}}$ parameters from a model fitted to the real UK data up to the first lockdown.

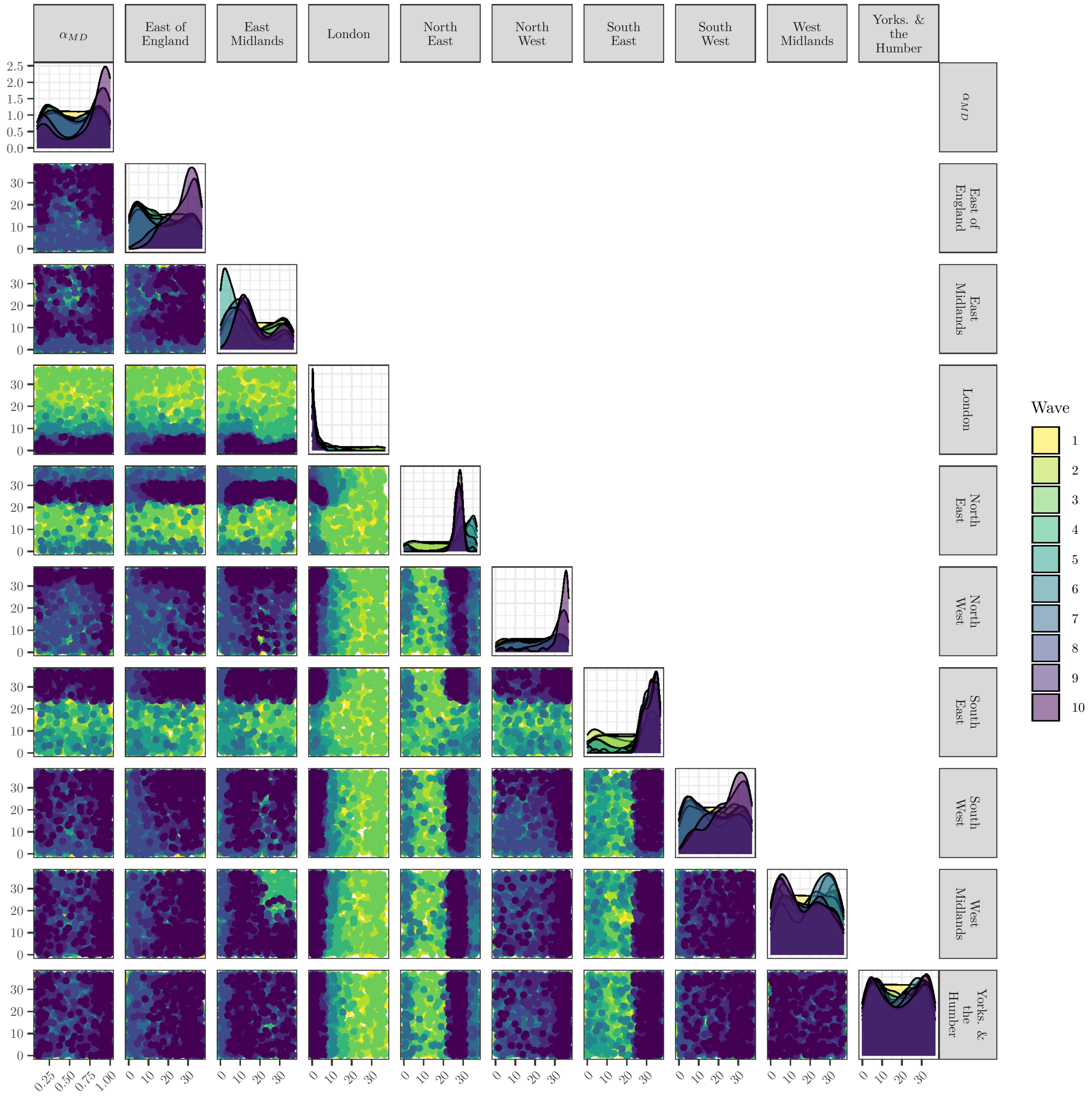


Figure S14: Inputs for Waves 1–10 for the $t_{r,\mathrm{MD}}$ parameters from a model fitted to the real UK data beyond the first lockdown.

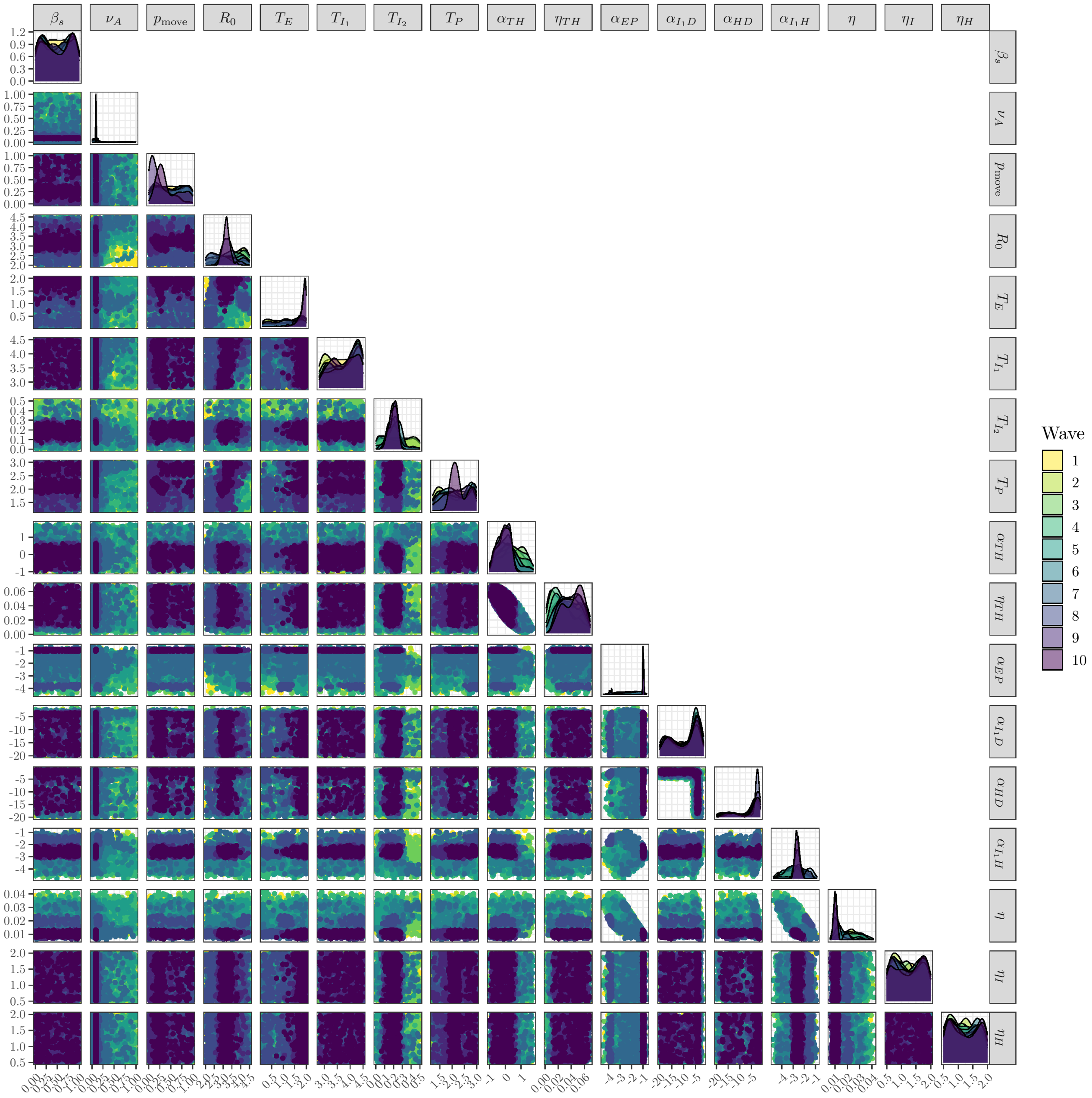


Figure S15: Inputs for Waves 1–10 for all parameters except the $t_{r,\text{MD}}$ parameters from a model fitted to the real UK data beyond the first lockdown.

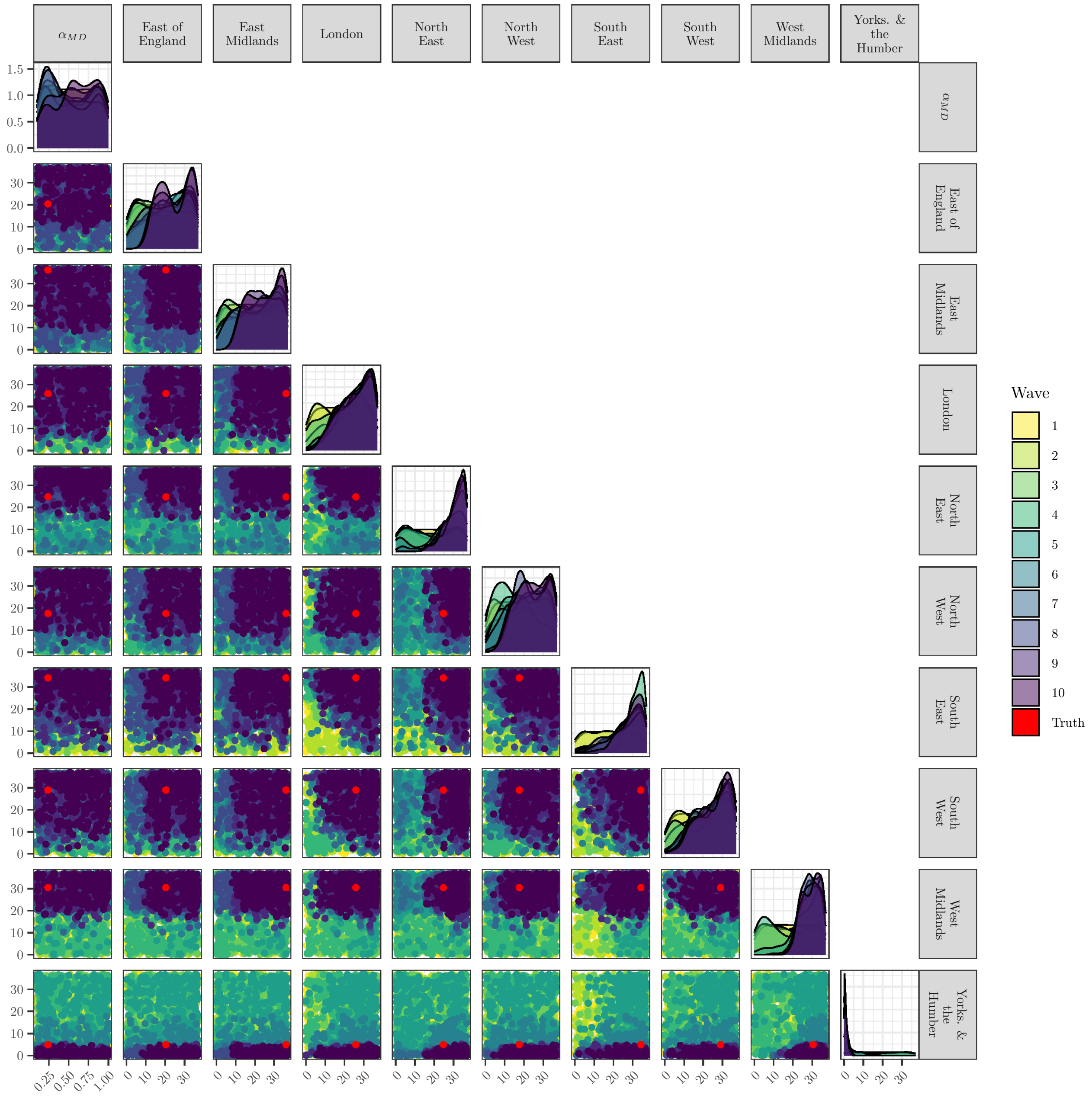


Figure S16: Inputs for Waves 1–10 for the $t_{r,\mathrm{MD}}$ parameters for a model fitted to the simulated data assuming data are available at the resolution of the model. The red points correspond to the true parameter values.

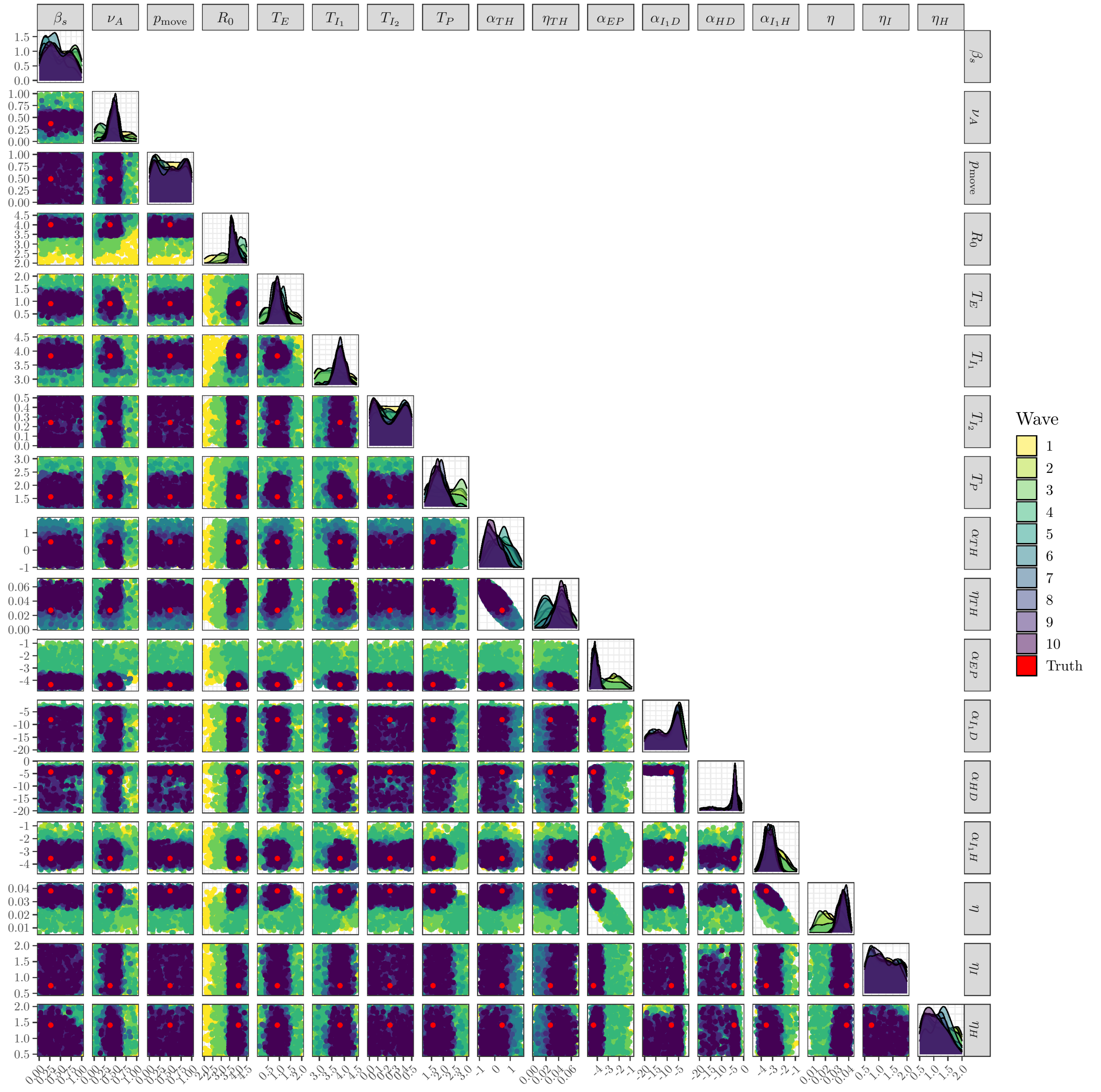


Figure S17: Inputs for Waves 1–10 for all parameters except the $t_{r,\text{MD}}$ parameters for a model fitted to the simulated data assuming data are available at the resolution of the model. The red points correspond to the true parameter values.
